# Effects of visual attention modulation on dynamic effective connectivity and visual fixation during own-face viewing in body dysmorphic disorder

**DOI:** 10.1101/2021.02.15.21249769

**Authors:** Wan-wa Wong, D. Rangaprakash, Joel P. Diaz-Fong, Natalie M. Rotstein, Gerhard S. Hellemann, Jamie D. Feusner

## Abstract

**Background:** In individuals with body dysmorphic disorder (BDD), selective attention biases and aberrant visual scanning patterns may cause imbalances in global vs. detailed visual processing, contributing to perceptual distortions for appearance. The mechanistic effects of modifying visual attention on brain function in BDD, which may be critical to developing perceptual-based treatments, have not been explored. This study tested the effects of visual-attention modulation on dorsal and ventral visual stream activation and connectivity, and eye behaviors.

**Methods:** We acquired functional magnetic resonance imaging data in 37 unmedicated adults with BDD and 30 controls. Participants viewed their faces under two conditions: a) unconstrained (naturalistically), and b) holding their gaze on the center of the image (visual-attention modulation), monitored with an eye-tracking camera. We analyzed activation and dynamic effective connectivity in dorsal and ventral visual streams and visual fixation duration.

**Results:** Visual-attention modulation resulted in longer fixation duration and reduced activation in dorsal and ventral visual streams in both groups compared with naturalistic viewing. Longer fixation duration was associated with greater effective connectivity from V1 to early dorsal visual stream during the second naturalistic viewing, across groups. During naturalistic viewing, there was greater V1 to early dorsal visual stream connectivity after, compared with before, visual-attention modulation.

**Conclusions:** When viewing one’s face, longer visual fixation may confer greater communication in dorsal visual system, facilitating global/holistic visual processing. The finding that reduction in visual scanning while viewing one’s face results in persistent effects during unconstrained viewing has implications for perceptual retraining treatment design for BDD.

## 1 Introduction

Body dysmorphic disorder (BDD) is marked by preoccupations with misperceived appearance defects, which sufferers believe render them ugly and deformed, and repetitive behaviors to check or fix one’s appearance. Commonly misperceived appearance features involve the face and head, although any body part can be of concern (1). The consequences can be profound, with high lifetime prevalence of suicide attempts (25%) (2) and hospitalization (50%) (3). 27 to 39 percent are delusional in their beliefs (4). BDD is still under-recognized, misdiagnosed, and understudied, although BDD has a high prevalence of ~2% in the general population (5). Some neurobiological models to explain vulnerability to BDD have been put forth (e.g., (6,7)) but a comprehensive understanding of this condition is still emerging.

Disturbances of visual information processing in BDD are likely critical neurobiological contributors to the core psychopathological feature of perceptual distortions of appearance (6,8). Our previous neuroimaging studies provide support for this premise. Using own-face (9), other-face (10), and house (11) stimuli as probes in functional magnetic resonance imaging (fMRI) studies, we found abnormally reduced activity in the dorsal visual stream (DVS) when viewing filtered images that contained only low spatial frequency information (i.e. conveying configural and holistic information). This led to the hypothesis that the hyper-scrutiny of miniscule appearance details could be mechanistically related to failing to “see” the appearance feature as an integrated whole, which may reflect an imbalance in global and local processing. This hypothesis has gained support from subsequent imaging and electro-cortical evidence (12,13). Adding to the hypothesized model, enhanced ventral visual stream (VVS) processing of high-detail images, and perception of faces as more unattractive when the magnitude of detailed processing increases, were found (12). Neuropsychological and psychophysical studies testing face and body inversion effects have corroborated the model of imbalance in global vs. local processing (14–19).

Further, selective attention biases potentially contribute to its psychopathological features (20). This could include aberrant patterns of visual attention, with excessive visual attention paid to perceived appearance defects, which is commonly observed phenomenologically (21). Studies using eye-tracking in BDD have found biased attention to facial areas deemed flawed, and a scanning pattern characterized by multiple fixations of brief duration (22,23).

In addition to psychophysical and visual task brain activation studies, functional connectivity (FC) studies have also been conducted in BDD (24,25). During an others’ face-viewing task, the BDD group demonstrated aberrant connectivity for low spatial frequency images within a face-processing network in the visual and temporal cortices, as well as between the fusiform face area and precuneus/posterior cingulate and insula (24). During a body-viewing task, individuals with BDD demonstrated reduced dorsal visual network connectivity (measured via independent component analysis) compared with healthy controls (25). These studies, testing face-processing and body-processing networks, resulted in findings consistent with a model of imbalances in global vs. local visual processing.

Given the phenomenology and the previous research in BDD, some current and proposed treatment approaches (26–28) incorporate visual-attention modifications. Yet, the neural mechanisms underlying aberrant visual attention and how the neurobiological substrates of potential targets are engaged by different visual-attention modification approaches are incompletely understood. A mechanistic understanding is critical for the development of, and ability to iteratively refine, effective clinical treatments.

We therefore designed an experiment to test the neurobiological mechanistic effects of a strategy of visual-attention modification (29). This strategy requires participants to visually fixate on a centered cross overlaid on their face photo, with eye-tracking camera monitoring. The purpose is to reduce visual scanning while viewing their face to enhance DVS activity, responsible for global/holistic visual processing, and to suppress VVS activity, responsible for detailed/analytic visual processing.

To examine directional connectivity, we employed dynamic effective connectivity (DEC) modeling (30) to assess connectivity changes from primary visual cortex (V1) to DVS and V1 to VVS over time in different viewing conditions (i.e. unconstrained “natural” viewing of their faces, NatV, and modulated viewing with fixation at a centered cross, ModV). In previous studies we found evidence of hypoactivation in early visual cortical areas such as V1 and early V2 for viewing own faces (9); as well as hypoactivation in later occipital (V2, V3) and parietal DVS regions, and hyperactivation in temporal fusiform VVS regions for viewing others’ faces (12). The primary goal was to investigate the effects of visual-attention modulation on the DVS and VVS activation and connectivity during own-face viewing within BDD and healthy controls. Secondarily, we explored if the modulation had differential effects between groups by comparing differences in brain activation, connectivity and visual fixation. We hypothesized increase fixation duration during ModV compared to NatV across groups, and that fixation duration would correlate with activation and connectivity in DVS. In addition, we hypothesized that ModV would enhance DVS and suppress VVS activation across groups. We also hypothesized increased connectivity from V1 to DVS and decreased connectivity from V1 to VVS during ModV compared to the first NatV in BDD and controls. Further, we hypothesized that during NatV after ModV there would be significant effects on DEC patterns within BDD and controls compared to the first NatV (i.e. a “carryover” effect of the ModV). Finally, we hypothesized that in BDD compared to controls there would be lower connectivity from V1 to DVS and higher connectivity from V1 to VVS during both ModV and NatV.

## 2 Materials and Methods

### 2.1 Participants

The UCLA Institutional Review Board approved the study. All participants provided informed written consent. Forty-three unmedicated adults with BDD and 35 healthy controls (CON) aged 18-40 years were recruited from the community and were enrolled. BDD participants met DSM-5 criteria for BDD, with face concerns. Those with concerns specifically about the region between their eyes were excluded due to the nature of ModV task. BDD participants could have comorbid depressive or anxiety disorders, since they commonly co-occur (See Supplementary Material S1 for exclusion criteria).

### 2.2 Clinical assessments

Eligibility was determined through telephone screening followed by a clinical interview with the study physician (JDF). The Mini International Neuropsychiatric Interview (MINI) and BDD Module (31,32) were administered. The Yale-Brown Obsessive-Compulsive Scale Modified for BDD (BDD-YBOCS) (33), Montgomery-Åsberg Depression Rating Scale (MADRS) (34), Brown Assessment of Beliefs Scale (BABS) (35), and the Hamilton Anxiety Scale (HAMA) (36) were administered (See Supplementary Material S2 for assessment details).

### 2.3 Task paradigm

There were two sets of stimuli for the NatV condition: photos of participant’s face and scrambled faces as the control task (Figure 1a). There were also two sets of stimuli for ModV condition: the same photos overlaid with a semi-transparent crosshair between the eyes, and the scrambled faces with a crosshair (Figure 1b).

**Figure 1.**
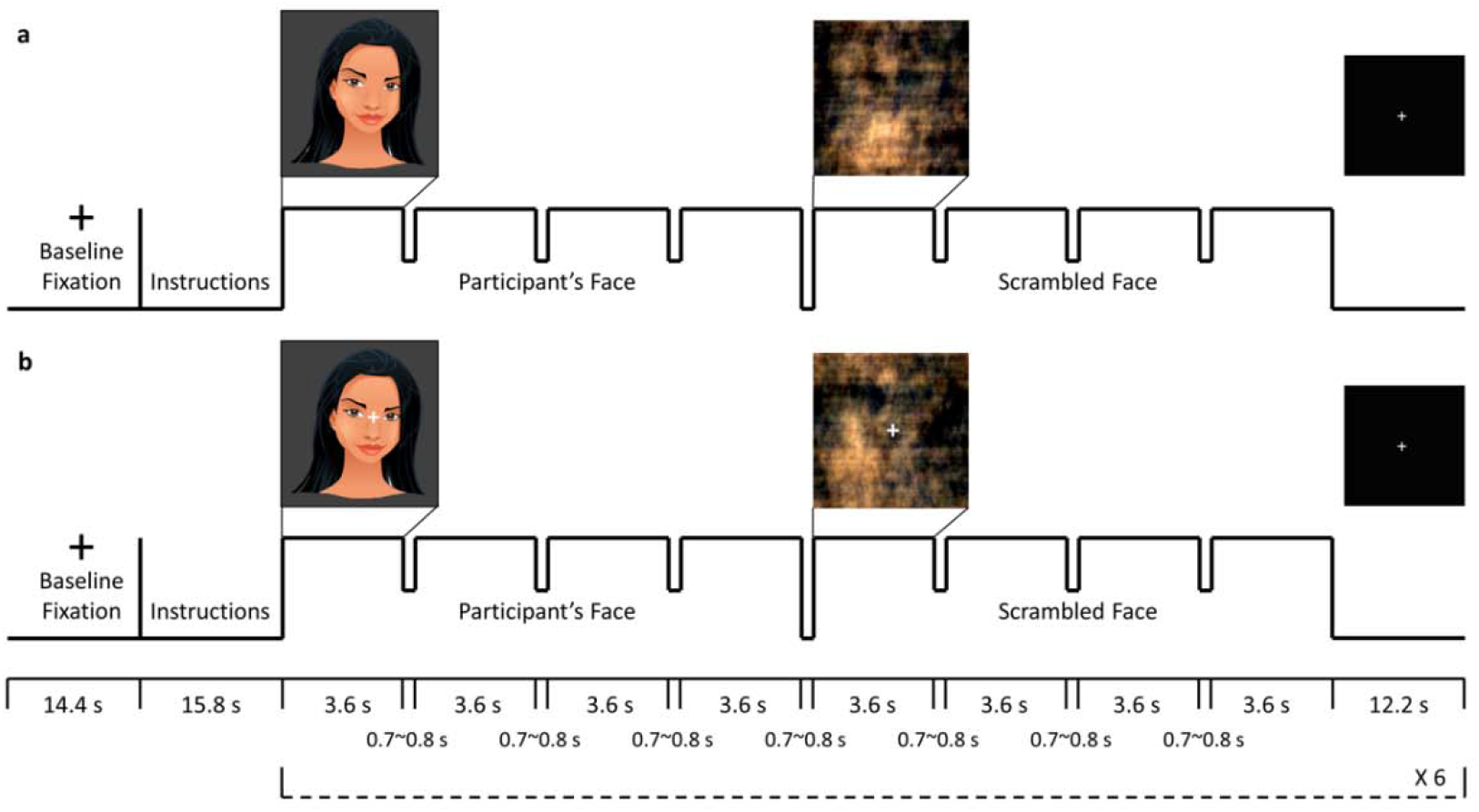
fMRI task paradigm. Four color photos of participants’ own faces at different, standardized angles were captured before the MRI session. A blocked design was used for the presentation of participant’s own face and scrambled face control stimuli for both (a) natural viewing and (b) visual modulation runs. The first 4 images were participant’s faces at different angles, and the next 4 images were scrambled faces. Each image was presented for 3.6 s, with a brief gap of 0.7~0.8 s for changing the image. A fixation with duration of 12.2 s was shown after the stimuli. The presentation of participant’s face and scrambled face stimuli was repeated six times in a single run. The stimuli for the visual modulation run (b) had a semi-transparent crosshair between the eyes of the participants’ faces and in the center of the scrambled faces. For the visual modulation run, participants were required to maintain their gaze on the crosshair. The rationale was that fixating visual gaze on the crosshair would reduce scanning associated with piecemeal/detailed processing and enhance holistic/global visual processing. To ensure task compliance for viewing the photos and crosshairs, gaze location was continuously monitored with the camera by the experimenters during the scan.

FMRI data were acquired while participants underwent two conditions. During NatV, participants were instructed to view the (unaltered) photos of their face and scrambled images of their face as they normally do. During ModV, they were instructed to view the same images while maintaining attention and eye gaze on the crosshair.

Participants were randomly assigned to one of the two counterbalanced groups for three fMRI runs: NatV-NatV-ModV (NNM) or NatV-ModV-NatV (NMN). They were instructed to press a button every time an image disappeared from the screen to ensure vigilance. Moreover, high-resolution MR-compatible display goggles (VisuaStimXGA, Resonance Technology, Inc.) with a right-side mounted infrared camera was used to present stimuli and record eye gaze. ViewPoint EyeTracker software (Arrington Research, Inc.) sampled pupil location at a rate of 30Hz. A 9-point calibration was used to normalize the eye gaze position relative to the screen. All values were normalized with respect to mapped x-axis and y-axis gaze values in a range of 0.0 to 1.0.

### 2.4 MRI data acquisition and preprocessing

MRI data were acquired on a 3T Siemens Prisma scanner. Data preprocessing was done using fMRIPrep 1.4.0 (37). See Supplementary Material S3-S5 for details of data acquisition and preprocessing, including quality control and motion correction.

### 2.5 Brain activation analysis

The preprocessed fMRI data were analyzed with a general linear model using FEAT in FSL 5.0.9 (38,39). For lower-level analysis, two different levels of contrasts (1^st^ level: unaltered face stimuli vs. fixation (baseline); 2^nd^ level: 1^st^ NatV vs. 2^nd^ NatV, 1^st^ NatV vs. ModV) were used. For higher-level analysis, group levels of contrasts (within-group level: BDD and CON; between-group level: BDD vs. CON) were used. We used DVS and VVS masks to test hypotheses regarding activation in these systems. See Supplementary Material S6 for more information.

### 2.6 Brain connectivity analysis

Fourteen regions-of-interest (ROIs) were derived from the Neurosynth (https://neurosynth.org/) functional meta-analysis in DVS and VVS (Figure 2). Blind-deconvolution (40) was performed on the timeseries extracted from these ROIs to minimize intra-subject variability in hemodynamic response function (HRF) (41) and to improve estimation of effective connectivity (42). DEC, a time-varying measure of directional connectivity between pairs of ROIs, was computed at each time point using time-varying Granger causality (GC) (30). The deconvolved timeseries were fitted into a dynamic multivariate autoregressive (dMVAR) model for estimating DEC between ROIs, which was solved in a Kalman-filter framework. The dMVAR model coefficients vary as a function of time, whose lengths were identical to the number of timepoints in the timeseries. See Supplementary Material S7 for more information. Twelve intra-hemispheric connections were chosen and divided into 4 categories: 1) VVS_Lower_ *(Calcarine to IOG)*, 2) VVS_Higher_ *(IOG to FG; IOG to ITG)*, 3) DVS_Lower_ *(Calcarine to SOG)*, and 4) DVS_Higher_ *(SOG to IPL; SOG to SPL)* (Figure 2). From these twelve connections, the timepoints associated with those trials of viewing unaltered faces were extracted for subsequent statistical analysis.

**Figure 2.**
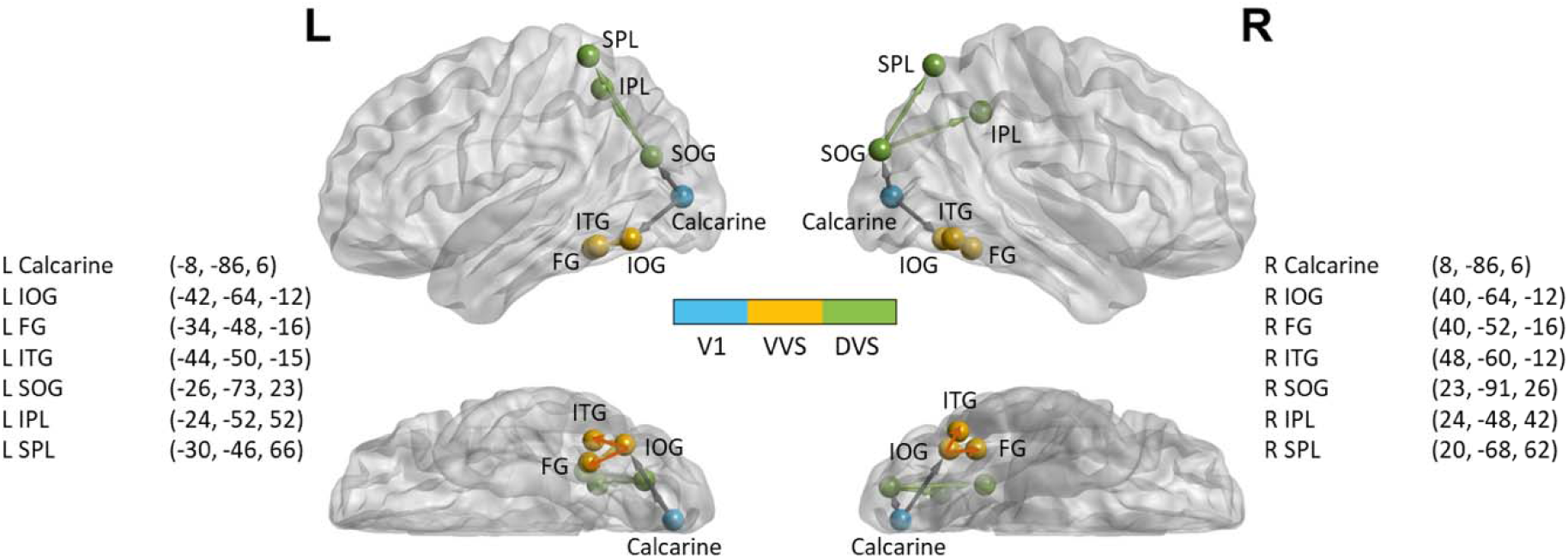
Locations of the 14 spherical ROIs used for dynamic effective connectivity analysis, overlaid on a brain surface with lateral and ventral views. The ROIs in the visual areas were defined using Neurosynth (https://neurosynth.org/) with the search terms including “primary visual”, “ventral visual”, “visual stream”, and “dorsal visual” to obtain maps generated with association tests. Clusters in the visual areas included 2 ROIs in V1 [bilateral calcarine], 6 ROIs in VVS [bilateral inferior occipital gyrus (IOG), fusiform gyrus (FG), and inferior temporal gyrus (ITG)], and 6 ROIs in DVS [bilateral superior occipital gyrus (SOG), inferior parietal lobul (IPL), and superior parietal lobule (SPL)]. The nomenclature is based on Eickhoff-Zilles macro labels from N27, implemented in AFNI. All spheres had a radius of 5 mm and the center-of-mass coordinates obtained from the clusters are x, y and z in the MNI space. This figure was prepared using BrainNet Viewer (43).

### 2.7 Gaze analysis

Pupil data were filtered with default settings in the ViewPoint software. Blinks were removed using a blink detection algorithm for low-speed eye-tracking (44). Missing values of less than four consecutive data points (~133ms) were linearly interpolated, to correct for flicker and loss of contact, considering that saccades typically take 100-130ms to program (45,46). Gaze position values were then smoothed using Savitzky-Golay filter (47), a simplified least square procedure which is suggested to perform well in low-speed eye-tracking that contains saccade amplitude greater than 5° (48). Fixations were identified using a velocity threshold algorithm (49) with a velocity threshold of 0.10°/s and a drift threshold of 0.30°/s. Fixations of less than 100ms were excluded from the analysis. Mean fixation duration was the main outcome variable to quantify fixation patterns when participants viewed their faces during the face stimuli.

### 2.8 Statistical analysis

Linear mixed models (LMM) were used to study whether the DEC was significantly influenced by experimental factors. Group (BDD or CON), order (NNM or NMN), run (1^st^ or 2^nd^ or 3^rd^ run), level (Lower or Higher), and their interactions were included in the model as fixed factors, with participant ID as random factor. Pairwise comparisons with Bonferroni correction (p<0.05) were performed afterwards to determine which factors significantly differed from each other. The LMM analysis was done for the separate DVS and VVS hypotheses. General linear model was used to analyze the mean fixation duration from eye-tracking data (within-subjects factor: run; between-subjects factors: group and order). If a significant three-way interaction effect was found, simple two-way interaction, simple-simple main effect, and simple-simple pairwise comparisons were computed post-hoc. Pearson correlation was used as exploratory follow-up analysis to determine associations between DEC, symptom severity measures of BDD-YBOCS and BABS, and mean fixation duration. Statistical tests were done using SPSS and R.

## 3 Results

### 3.1 Sample characteristics

Forty-three BDD participants and 35 CON were eligible and scanned. Among these, we excluded one BDD and one CON due to task noncompliance, four BDD and four CON due to excessive motion artifacts, and one BDD due to fMRIPrep errors. Thirty-seven BDD and 30 CON were finally included in the subsequent analyses (Table 1).

**Table 1.**
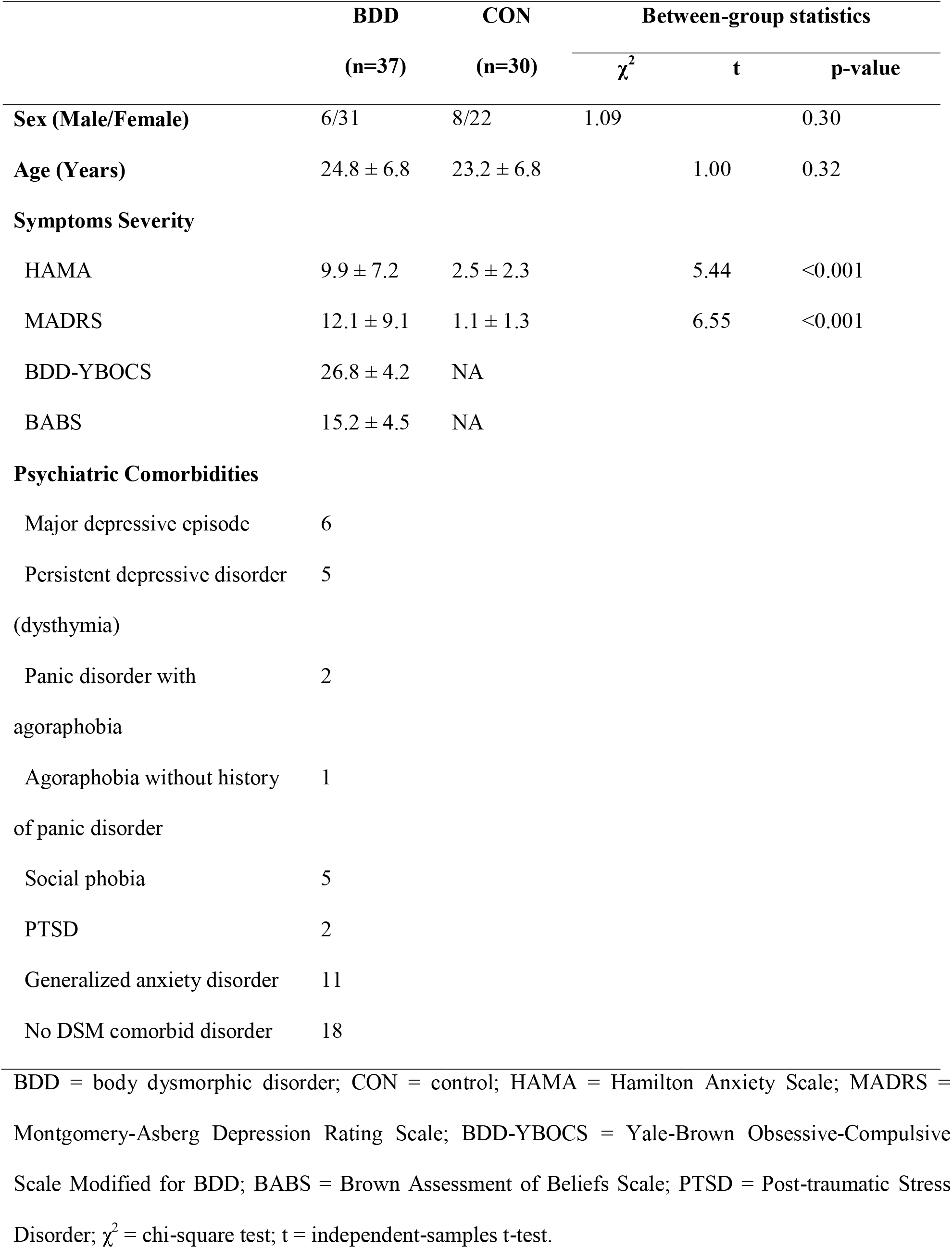
Sample characteristics

|  | **BDD**<br>**(n=37)** | **CON**<br>**(n=30)** | **Between-group statistics** |  |  |
| --- | --- | --- | --- | --- | --- |
| | | | $\chi^2$ | t | p-value |
| **Sex (Male/Female)** | 6/31 | 8/22 | 1.09 |  | 0.30 |
| **Age (Years)** | 24.8 ± 6.8 | 23.2 ± 6.8 |  | 1.00 | 0.32 |
| **Symptoms Severity** |  |  |  |  |  |
| HAMA | 9.9 ± 7.2 | 2.5 ± 2.3 |  | 5.44 | <0.001 |
| MADRS | 12.1 ± 9.1 | 1.1 ± 1.3 |  | 6.55 | <0.001 |
| BDD-YBOCS | 26.8 ± 4.2 | NA |  |  |  |
| BABS | 15.2 ± 4.5 | NA |  |  |  |
| **Psychiatric Comorbidities** |  |  |  |  |  |
| Major depressive episode | 6 |  |  |  |  |
| Persistent depressive disorder<br>(dysthymia) | 5 |  |  |  |  |
| Panic disorder with<br>agoraphobia | 2 |  |  |  |  |
| Agoraphobia without history<br>of panic disorder | 1 |  |  |  |  |
| Social phobia | 5 |  |  |  |  |
| PTSD | 2 |  |  |  |  |
| Generalized anxiety disorder | 11 |  |  |  |  |
| No DSM comorbid disorder | 18 |  |  |  |  |

BDD = body dysmorphic disorder; CON = control; HAMA = Hamilton Anxiety Scale; MADRS = Montgomery-Asberg Depression Rating Scale; BDD-YBOCS = Yale-Brown Obsessive-Compulsive Scale Modified for BDD; BABS = Brown Assessment of Beliefs Scale; PTSD = Post-traumatic Stress Disorder; $\chi^2$ = chi-square test; t = independent-samples t-test.

### 3.2 Gaze patterns

Nine participants were excluded for the eye-tracking analysis: three BDD due to device malfunction, two BDD due to noncompliance, and two BDD and two CON due to poor data quality. A three-way mixed ANOVA was performed to evaluate the effects of group, order and run on the mean fixation duration. There was no significant three-way interaction between group, order and run, F(1.74,83.62)=2.27, p=0.12. There was significant two-way interaction between order and run, F(1.74,83.62)=16.03, p<0.001. Following this up by a simple main effect analysis with a Bonferroni adjustment (statistical significance being accepted at p<0.025), there were significant simple main effects of run on the mean fixation duration for the NMN order, F(1.57,36.2)=7.74, p=0.003, and for the NNM order, F(1.44,38.9)=11.9, p=0.0004. Simple pairwise comparisons were computed between runs for the NMN and NNM orders, with a Bonferroni adjustment. The mean fixation duration was significantly different between the 1^st^ and 3^rd^ runs (ModV > 1^st^ NatV, p=0.023), and between the 2^nd^ and 3^rd^ runs (ModV > 2^nd^ NatV, p=0.014) for the NNM order. There was a trend between the 2^nd^ and 3^rd^ runs (ModV > 2^nd^ NatV, p=0.065) for the NMN order (Figure 3). In sum, across groups and for both NNM and NMN orders, as expected the mean fixation duration lasted longer during ModV compared to NatV.

**Figure 3.**
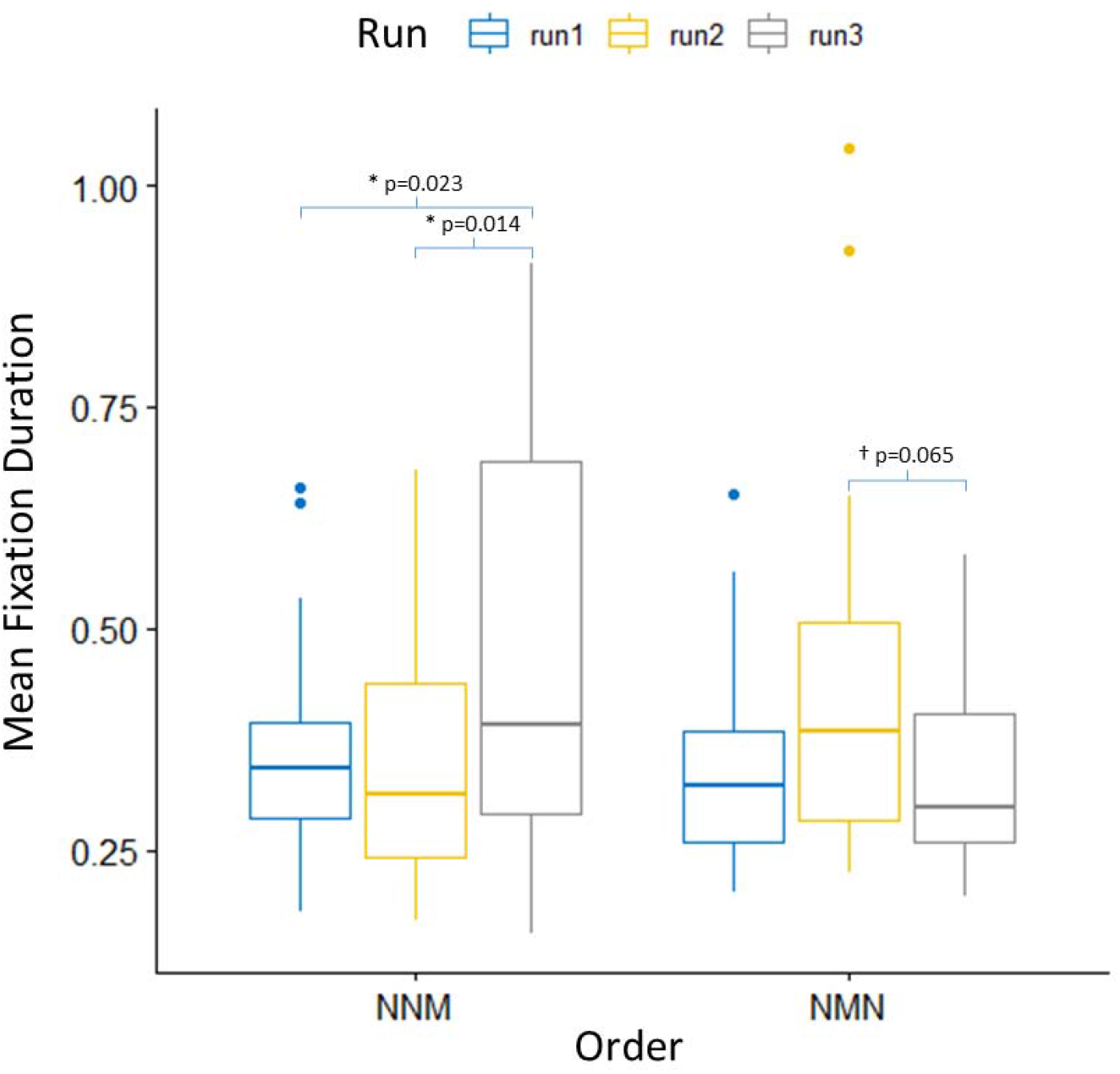
Mean fixation duration across different orders and runs. Results are shown after collapsing the group factor, as BDD and CON, had a common pattern of fixation duration across the three runs for each order.

### 3.3 Brain activation patterns

Activation in the DVS and VVS in response to unaltered face stimuli relative to baseline was greater during 1^st^ NatV compared with ModV for both BDD and CON (Figure S1). No significant difference was found in the activation in response to unaltered face stimuli compared with baseline for the contrast of 1^st^ NatV vs. 2^nd^ NatV, in BDD or CON. There was no significant difference for any contrasts between BDD and CON. There were no significant associations between activation and mean fixation duration for any condition, in BDD and CON.

### 3.4 Brain connectivity patterns

In the DVS, there were significant effects for ‘Level’ (p<0.001), ‘Order x Run’ (p<0.001), ‘Run x Level’ (p<0.001), ‘Group x Order x Run’ (p=0.001), ‘Group x Order x Level’ (p<0.001), ‘Order x Run x Level’ (p<0.001), and ‘Group x Order x Run x Level’ (p=0.018). Pairwise comparisons were computed between different runs, with a Bonferroni adjustment. For DVS_Lower_, both BDD and CON with the NMN order showed greater DEC during 2^nd^ NatV compared to 1^st^ NatV and ModV (Figure 4), while both BDD and CON with the NNM order exhibited greater DEC during 1^st^ NatV compared to 2^nd^ NatV and ModV (Figure S2a). For DVS_Higher_, BDD with the NMN order showed greater DEC during ModV and 2^nd^ NatV compared to 1^st^ NatV, while BDD with the NNM order only showed greater DEC during 2^nd^ NatV compared to 1^st^ NatV. However, CON with the NNM order showed greater DEC during 2^nd^ NatV and ModV compared to 1^st^ NatV, while CON with the NMN order only showed greater DEC during ModV compared to 1^st^ NatV (Figure S2a). All these differences were significant at p<0.05, corrected.

**Figure 4.**
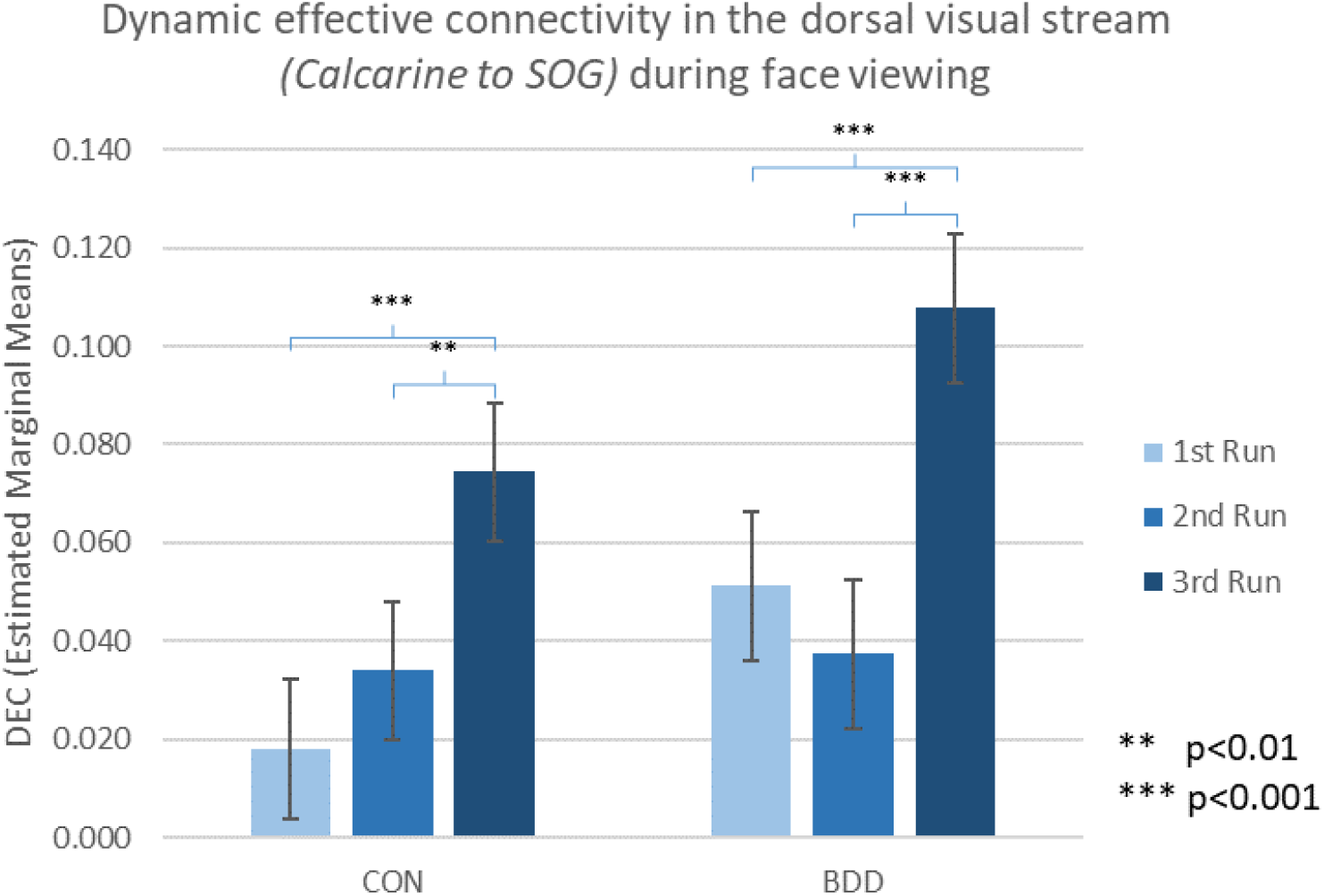
Estimated marginal means of dynamic effective connectivity for the dorsal visual steam (lower connections, bilateral V1 to superior occipital gyrus - SOG) across CON and BDD groups with the NMN order. The participants randomized to the NMN order received natural viewing (N), modulated viewing (M), and then natural viewing (N) as the 1^st^, 2^nd^ and 3^rd^ runs. The p-values were Bonferroni corrected.

In the VVS, there were significant effects for ‘Run’ (p<0.001), ‘Level’ (p<0.001), ‘Order x Run’ (p<0.001), ‘Run x Level’ (p<0.001), ‘Group x Order x Run’ (p<0.001), ‘Group x Order x Level’ (p=0.050), ‘Group x Run x Level’ (p<0.001), ‘Order x Run x Level’ (p=0.017), and ‘Group x Order x Run x Level’ (p<0.001). From pairwise comparisons between different runs (p<0.05, Bonferroni corrected), for both VVS_Lower_ and VVS_Higher_, participants with the NNM order showed greater DEC during 1^st^ NatV compared to 2^nd^ NatV. There was no common pattern between BDD and CON with NMN order (Figure S2b).

### 3.5 Relationships between brain connectivity, visual fixation and clinical symptoms

For DVS_Lower_, participants with the NNM order exhibited greater DEC during 1^st^ NatV compared with the 2^nd^ NatV and ModV, while participants with the NMN order showed greater DEC during the 2^nd^ NatV compared to the 1^st^ NatV and ModV. Since a common pattern of DEC changes across the 3 runs was discovered for DVS_Lower_ from the results of BDD and CON, the inter-relationships between DEC of DVS_Lower_, visual fixation duration and clinical symptoms (BDD-YBOCS and BABS) were further explored with post hoc tests. Across groups, mean fixation duration positively correlated with DEC during the 2^nd^ NatV for DVS_Lower_ (r=0.280, p=0.04) (Figure 5); those with shorter fixation duration had weaker DEC for DVS_Lower_ during 2^nd^ NatV. Negative trends were observed between BDD-YBOCS and mean fixation duration during 1^st^ and 2^nd^ NatV in BDD (1^st^ NatV: r=-0.301, p=0.113; 2^nd^ NatV: r=-0.342, p=0.070) (Figure 6); BDD individuals with more severe BDD symptom tended to have shorter fixation duration during NatV.

**Figure 5.**
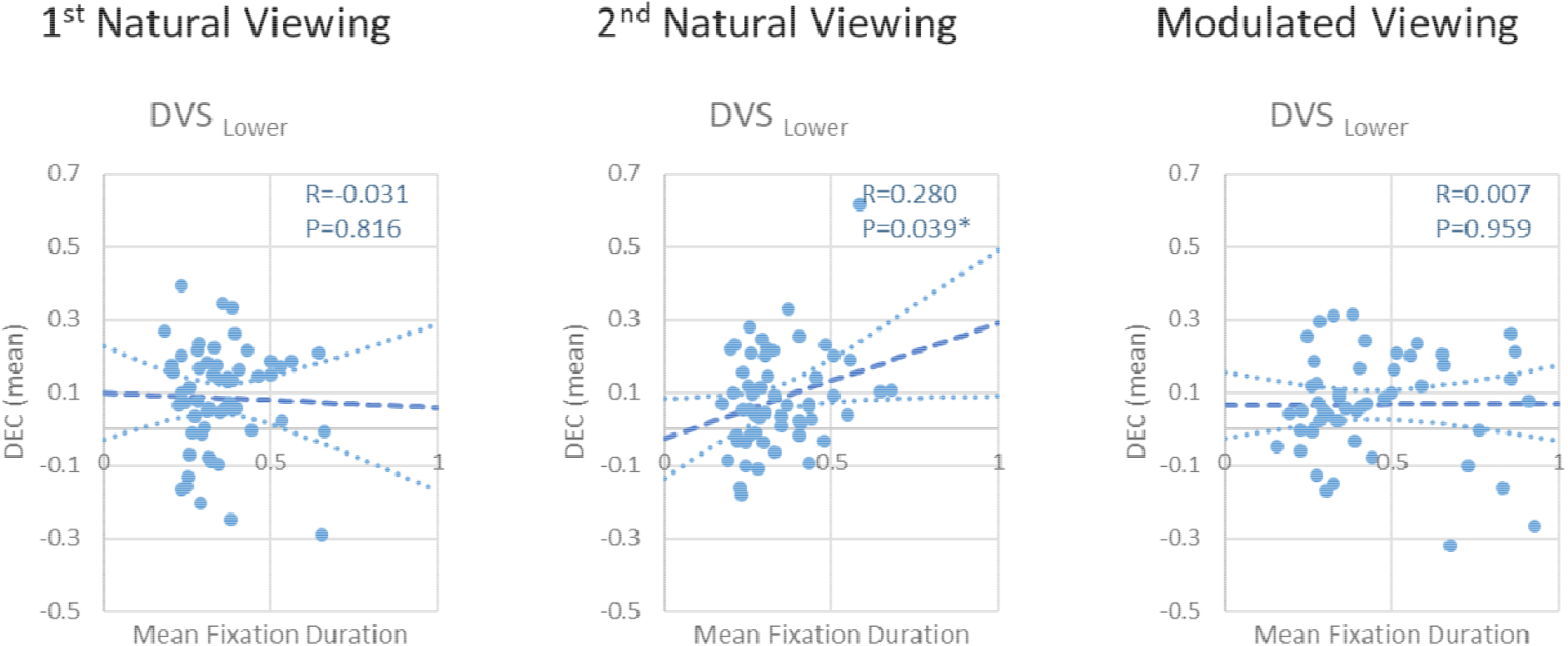
Correlations between mean dynamic effective connectivity (DEC) and mean fixation duration across all participants during 1^st^ NatV, 2^nd^ NatV, and ModV face viewing.

**Figure 6.**
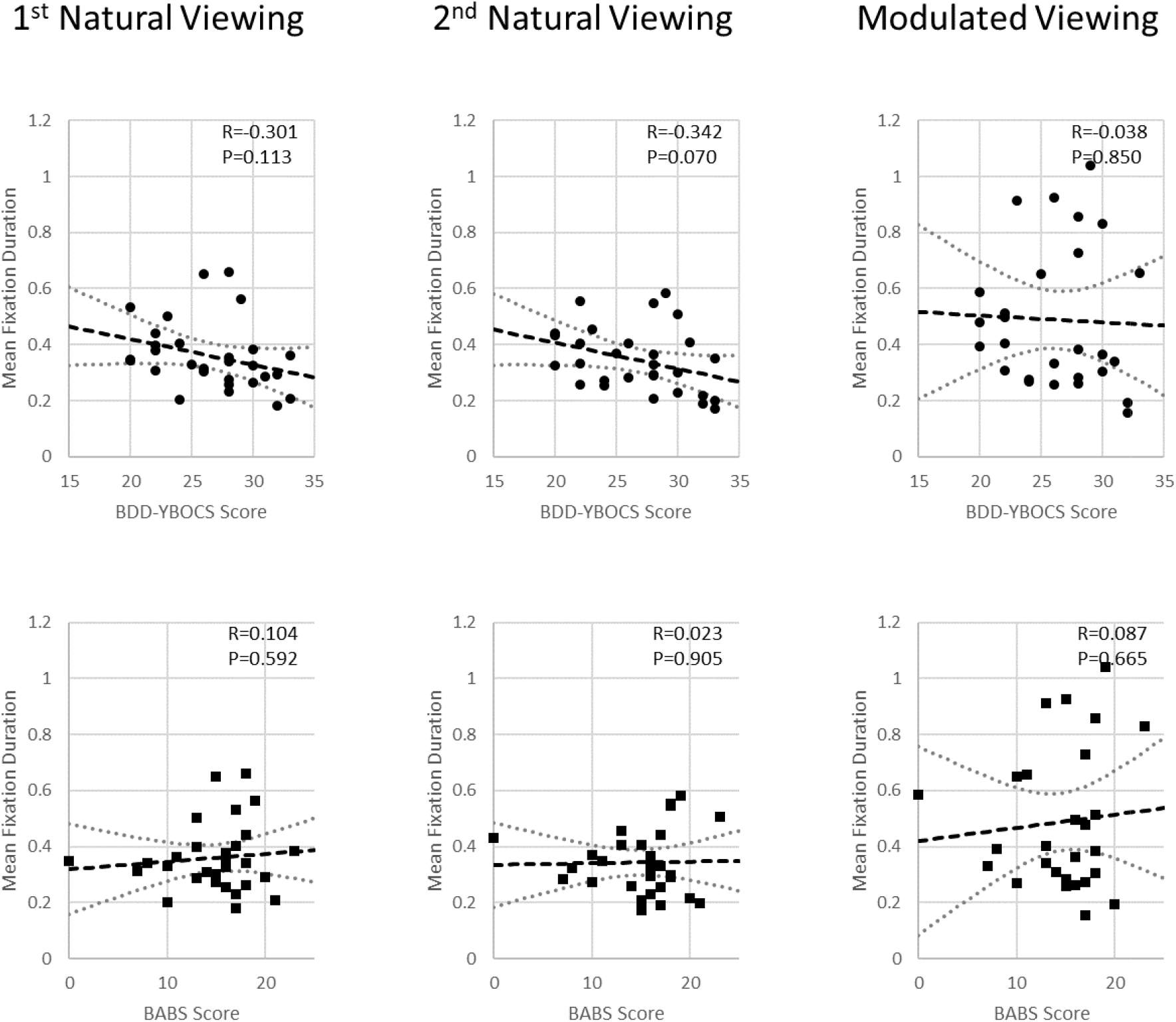
Correlations between mean fixation duration and BDD-YBOCS scores and BABS scores across BDD participants during 1^st^ NatV, 2^nd^ NatV, and ModV.

## 4 Discussion

The goal of this study was to understand how brain activation, connectivity, and visual fixation patterns when viewing one’s face – the primary area of appearance concern for most with BDD – change under conditions of modulated visual attention. We specifically investigated a) how brain activation, connectivity, and visual fixation are influenced by visual-attention modulation, b) if there are subsequent “carryover” effects when viewing one’s face naturalistically after visual-attention modulation, and c) whether these effects differ in individuals with BDD compared with CON. Visual-attention modulation resulted in longer fixation duration and reduced activation in DVS and VVS in BDD and CON compared with naturalistic viewing. Across groups, longer fixation duration was associated with greater dynamic effective connectivity from V1 to early DVS during the second naturalistic viewing. There was stronger connectivity from V1 to early DVS during the second naturalistic viewing following visual-attention modulation, compared with the corresponding first naturalistic viewing. Those with more severe BDD symptoms showed a trend for shorter fixation duration during naturalistic viewing. These findings shed light on the inter-relationships between brain connectivity, eye behaviors and BDD symptoms. Importantly, they demonstrate the mechanistic effects of a brief attention modulation intervention of holding gaze constant on brain connectivity and visual fixation, which may have implications for novel perceptual retraining treatment designs.

BDD and CON showed longer fixation duration during ModV compared with NatV. This was expected due to the task nature that required them to fixate their gaze on a centered cross. It also demonstrated overall compliance with the ModV instructions. No significant associations were found between mean fixation duration and clinical scores during ModV. This suggests that, invariant to symptom severity, BDD participants were able to visually fixate on the crosshair, which is an area of their face for which they did not have significant appearance concerns (only one potential BDD participant was deemed ineligible due to having a concern for the area corresponding to the fixation cross). Yet, there were trends for negative associations between mean fixation duration and BDD-YBOCS scores during both NatV conditions; those with more severe BDD symptoms had shorter fixation duration during NatV, a trend that was stronger during the second NatV. Previous eye-tracking studies in BDD have shown aberrant scan-paths when viewing stimuli such as faces. These scan-paths are generally characterized by either a “focused” pattern – paying attention to areas of concern, or an “avoidant” pattern – avoiding perceived defects (22,23,50,51). In these studies, BDD participants showed aberrant eye movements, including enhanced selective visual attention to imagined defects, overfocus on negative attributes, or atypical scanning behaviors with more blinks, fewer fixations, and less visual attention paid to prominent facial features. Abnormalities in face-processing seem particularly evident in BDD when viewing own-faces and faces showing negative or neutral emotional expressions (50,51). In a study examining attention to attractive vs. unattractive parts of one’s own and other’s faces in participants with BDD, bulimia nervosa (BN), and CON, BDD and BN participants spent less time looking at attractive parts of their *own* face than CON, yet more time looking at attractive parts than unattractive parts of *other’s* faces (52). This suggests ignoring of positive aspects of one’s face in BDD, and/or upward social comparison, either or both of which could account for the increase in negative emotions observed in BDD after face viewing. In the current study, shorter fixation duration during NatV, in those with more severe BDD symptoms, suggests multiple short-duration fixation patterns interspersed with an increased number of saccades for scanning multiple facial details. This could reflect heightened attention to multiple perceived appearance flaws, or, alternatively, an unwillingness to fixate on any one area of their own faces more than briefly due to a triggering of negative emotions.

Another important finding was a positive association between fixation duration and connectivity from V1 to SOG during the second NatV, across participants. Thus, those with shorter fixation duration had weaker connectivity during the second NatV. In general, eye-movement parameters, including fixation duration and saccade amplitude, can be used to characterize distinct modes of visual processing (53), which may indicate differential involvement of dorsal and ventral systems in saccade planning and information processing. Although we did not directly quantify saccades (due to limitations of the data from the google-mounted eye-tracker camera), fixation on a crosshair would be expected to be accompanied by fewer saccades than naturalistic viewing.

Saccades have been found to suppress low spatial frequency (dorsal pathway) contrast sensitivity (54), suggesting a reduction of global/configural processing. Moreover, the frontal eye fields for controlling visual attention and eye movements have dense connections with the occipitoparietal network (DVS) (55), such that reduction of eye scanning (also reduced occurrence of saccades) would enhance DVS activity. Our findings corroborate this model, that longer fixation duration associates with stronger effective connectivity in the early DVS. Thus, potential changes in attentional allocation in conjunction with eye gaze behavior may have a modulatory effect on the early DVS, especially in later periods of face viewing that were evident during the second NatV. Alternatively, previous studies of eye behaviors describe a “pre-attentive” mode, in which scanning eye movements are predominant with brief fixations and large saccades, while in an “attentive” mode, long fixations and small saccades are present, leading to detailed inspection (53,56,57). In theory, pre-attentive scanning behavior could reflect dorsal pathway processing, while attentive inspection behavior could reflect ventral pathway processing (53,58). However, it is important to note that the studies characterizing these viewing modes were based on scene viewing and may not apply to face processing, for which humans have high expertise and specialized visual “templates,” and did not specifically examine dynamic connectivity patterns as in the current study.

Contrary to our hypothesis, there were no significant group differences in connectivity from V1 to DVS or V1 to VVS during the ModV or NatV conditions. Although, during the first NatV, the magnitude of connectivity was slightly lower in early DVS, and slightly higher in early and later VVS connectivity in BDD than CON (Figure S3). As the experiment was designed to test within-subject effects of ModV as the primary goal, it is possible that the study was underpowered to detect group differences in DEC owing to the number of main effects and interactions.

There are several important implications of these findings that could impact future translational research. These results provide early evidence that changing eye-gaze behaviors might change the balance of global vs. local processing mediated by the DVS. This has been suggested in other ongoing (59) or planned (60) treatment protocols. The observation of increased DVS connectivity during the NatV following the brief period of ModV (fixating on a non-concerning region of the face) suggests the possibility of a persistent DVS effect that may enhance global/holistic processing. A similar phenomenon was demonstrated in a study in which exposure to a Navon visual stimulus (61) – a large letter made of smaller letters that was presented in a way to promote a global bias – induced global processing and temporarily reversed visual processing biases in individuals with great body image concerns (62).

In this study, although there was no significant increase in visual fixation duration from ModV to the following NatV, the DVS connectivity magnitude nevertheless scaled with the fixation duration during this period. Thus, individuals with longer fixation duration, which may have persisted from the preceding attention modulation, could have experienced enhanced DVS connectivity. In this second NatV, there were no explicit instructions other than to view one’s face naturalistically, so changes in gaze patterns were likely implicit, although some participants might have willfully tried to reduce scanning during this period.

There are several limitations to consider. The study population underrepresents the proportion of males with BDD in the general population (63,64), thus findings may not generalize. Another limitation is that we did not assess participants’ emotional states during face viewing (in the interest of not interrupting natural processes involved in face viewing that might be disrupted by self-reflection). Thus, we could not investigate how degree of emotional arousal, such as anxiety (65), affects visual system activity. Moreover, we were unable to use areas-of-interest on the face photographs due to limitations in positional stability of the goggle-mounted eye-tracking camera that otherwise might be informative about viewing patterns of areas of concern during NatV after ModV.

In conclusion, these findings provide evidence of enhanced dynamic connectivity from V1 to early DVS when viewing faces naturalistically after visual-attention modulation. This suggests that visual-attention modulation may have a subsequent carryover effect on the connectivity patterns during natural viewing of one’s face afterwards, which could enhance global/configural visual processing. The clinical relevance is underscored by the observation that those with more severe BDD symptoms had shorter fixation duration, and that longer visual fixation duration associated with greater DVS connectivity. Visual-attention modulation thus holds promise for future studies of perceptual retraining for BDD.

## Data Availability

The National Institute of Health (NIH) and the National Institute of Mental Health (NIMH) have developed a federation of data repositories to store the collection of data from participants in research studies related to mental health. The information collected from our study "Understanding the dynamics of visual processing abnormalities in body dysmorphic disorder" is stored in the Research Domain Criteria Database (RDoCdb).

https://nda.nih.gov/edit_collection.html?id=2652

## Acknowledgments

The authors declare no conflicts of interest.

This study was supported by the National Institute of Mental Health (R21MH110865 to JDF, R01MH121520 to JDF), and the Nathan Cumming Foundation (JDF).

## Author Contributions

WW, RD, JD, and NR were responsible for data analysis and paper writing. GH was responsible for statistical modeling. JDF was responsible for clinical assessment, experimental design, and paper writing. All authors read and approved the submitted manuscript.

## References

1. Phillips KA (2005): The Broken Mirror. New York, NY: Oxford University Press.

2. Phillips KA, Menard W (2006): Suicidality in body dysmorphic disorder: a prospective study. Am J Psychiatry163: 1280–1282.

3. Phillips KA, McElroy SL, Keck PE Jr, Hudson JI, Pope HG Jr (1994): A comparison of delusional and nondelusional body dysmorphic disorder in 100 cases. Psychopharmacol Bull30: 179–186.

4. Phillips KA (2004): Psychosis in body dysmorphic disorder. J Psychiatr Res 38: 63–72.

5. Veale D, Gledhill LJ, Christodoulou P, Hodsoll J (2016): Body dysmorphic disorder in different settings: A systematic review and estimated weighted prevalence. Body Image 18: 168–186.

6. Li W, Arienzo D, Feusner JD (2013): Body Dysmorphic Disorder: Neurobiological Features and an Updated Model. Zeitschrift Für Klinische Psychologie Und Psychotherapie, vol. 42. pp 184–191.

7. Grace SA, Labuschagne I, Kaplan RA, Rossell SL (2017): The neurobiology of body dysmorphic disorder: A systematic review and theoretical model. Neuroscience & Biobehavioral Reviews, vol. 83. pp 83–96.

8. Beilharz F, Castle DJ, Grace S, Rossell SL (2017): A systematic review of visual processing and associated treatments in body dysmorphic disorder. Acta Psychiatr Scand 136: 16–36.

9. Feusner JD, Moody T, Hembacher E, Townsend J, McKinley M, Moller H, Bookheimer S (2010): Abnormalities of visual processing and frontostriatal systems in body dysmorphic disorder. Arch Gen Psychiatry 67: 197–205.

10. Feusner JD, Townsend J, Bystritsky A, Bookheimer S (2007): Visual information processing of faces in body dysmorphic disorder. Arch Gen Psychiatry 64: 1417–1425.

11. Feusner JD, Hembacher E, Moller H, Moody TD (2011): Abnormalities of object visual processing in body dysmorphic disorder. Psychol Med 41: 2385–2397.

12. Li W, Lai TM, Bohon C, Loo SK, McCurdy D, Strober M, et al. (2015): Anorexia nervosa and body dysmorphic disorder are associated with abnormalities in processing visual information. Psychol Med 45: 2111–2122.

13. Li W, Lai TM, Loo SK, Strober M, Mohammad-Rezazadeh I, Khalsa S, Feusner J (2015): Aberrant early visual neural activity and brain-behavior relationships in anorexia nervosa and body dysmorphic disorder. Front Hum Neurosci 9: 301.

14. Deckersbach T, Savage CR, Phillips KA, Wilhelm S, Buhlmann U, Rauch SL, et al. (2000): Characteristics of memory dysfunction in body dysmorphic disorder. J Int Neuropsychol Soc 6: 673–681.

15. Feusner JD, Moller H, Altstein L, Sugar C, Bookheimer S, Yoon J, Hembacher E (2010): Inverted face processing in body dysmorphic disorder. J Psychiatr Res 44: 1088–1094.

16. Jefferies K, Laws KR, Fineberg NA (2012): Superior face recognition in Body Dysmorphic Disorder. J Obsessive Compuls Relat Disord 1: 175–179.

17. Stangier U, Adam-Schwebe S, Müller T, Wolter M (2008): Discrimination of facial appearance stimuli in body dysmorphic disorder. J Abnorm Psychol 117: 435–443.

18. Mundy E M, Sadusky A (2014): Abnormalities in visual processing amongst students with body image concerns. Adv Cogn Psychol 10: 39–48.

19. Dhir S, Ryan HS, McKay EL, Mundy ME (2018): Parameters of visual processing abnormalities in adults with body image concerns. PLoS One 13: e0207585.

20. Johnson S, Williamson P, Wade TD (2018): A systematic review and meta-analysis of cognitive processing deficits associated with body dysmorphic disorder. Behav Res Ther 107: 83–94.

21. Phillips KA (2009): Understanding Body Dysmorphic Disorder. New York, NY: Oxford University Press.

22. Greenberg JL, Reuman L, Hartmann AS, Kasarskis I, Wilhelm S (2014): Visual hot spots: an eye tracking study of attention bias in body dysmorphic disorder. J Psychiatr Res 57: 125–132.

23. Grocholewski A, Kliem S, Heinrichs N (2012): Selective attention to imagined facial ugliness is specific to body dysmorphic disorder. Body Image 9: 261–269.

24. Moody TD, Sasaki MA, Bohon C, Strober MA, Bookheimer SY, Sheen CL, Feusner JD (2015): Functional connectivity for face processing in individuals with body dysmorphic disorder and anorexia nervosa. Psychol Med 45: 3491–3503.

25. Moody TD, Morfini F, Cheng G, Sheen CL, Kerr WT, Strober M, Feusner JD (2020): Brain activation and connectivity in anorexia nervosa and body dysmorphic disorder when viewing bodies: relationships to clinical symptoms and perception of appearance. Brain Imaging Behav. https://doi.org/10.1007/s11682-020-00323-5

26. Wilhelm S, Phillips KA, Greenberg JL, O’Keefe SM, Hoeppner SS, Keshaviah A, et al. (2019): Efficacy and posttreatment effects of therapist-delivered cognitive behavioral therapy vs supportive psychotherapy for adults with body dysmorphic disorder: A randomized clinical trial. JAMA Psychiatry 76: 363–373.

27. Johnson S, Egan SJ, Andersson G, Carlbring P, Shafran R, Wade TD (2019): Internet-delivered cognitive behavioural therapy for perfectionism: Targeting dysmorphic concern. Body Image 30: 44–55.

28. Beilharz F, Rossell SL (2017): Treatment Modifications and Suggestions to Address Visual Abnormalities in Body Dysmorphic Disorder. J Cogn Psychother 31: 272–284.

29. Feusner J, Deshpande R, Strober M (2017): A translational neuroscience approach to body image disturbance and its remediation in anorexia nervosa. Int J Eat Disord 50: 1014–1017.

30. Büchel C, Friston KJ (1998): Dynamic changes in effective connectivity characterized by variable parameter regression and Kalman filtering. Hum Brain Mapp 6: 403–408.

31. Eisen JL, Phillips KA, Coles ME, Rasmussen SA (2004): Insight in obsessive compulsive disorder and body dysmorphic disorder. Compr Psychiatry 45: 10–15.

32. Rief W, Buhlmann U, Wilhelm S, Borkenhagen A, Brähler E (2006): The prevalence of body dysmorphic disorder: a population-based survey. Psychol Med 36: 877–885.

33. Phillips KA, Hollander E, Rasmussen SA, Aronowitz BR, DeCaria C, Goodman WK (1997): A severity rating scale for body dysmorphic disorder: development, reliability, and validity of a modified version of the Yale-Brown Obsessive Compulsive Scale. Psychopharmacol Bull 33: 17– 22.

34. Montgomery SA, Asberg M (1979): A new depression scale designed to be sensitive to change. Br J Psychiatry 134: 382–389.

35. Eisen JL, Phillips KA, Baer L, Beer DA, Atala KD, Rasmussen SA (1998): The Brown Assessment of Beliefs Scale: reliability and validity. Am J Psychiatry 155: 102–108.

36. Hamilton M (1959): The assessment of anxiety states by rating. Br J Med Psychol 32: 50–55.

37. Esteban O, Markiewicz CJ, Blair RW, Moodie CA, Isik AI, Erramuzpe A, et al. (2019): fMRIPrep: a robust preprocessing pipeline for functional MRI. Nat Methods 16: 111–116.

38. Woolrich MW, Ripley BD, Brady M, Smith SM (2001): Temporal autocorrelation in univariate linear modeling of FMRI data. Neuroimage 14: 1370–1386.

39. Woolrich MW, Behrens TEJ, Beckmann CF, Jenkinson M, Smith SM (2004): Multilevel linear modelling for FMRI group analysis using Bayesian inference. Neuroimage 21: 1732–1747.

40. Wu G-R, Liao W, Stramaglia S, Ding J-R, Chen H, Marinazzo D (2013): A blind deconvolution approach to recover effective connectivity brain networks from resting state fMRI data. Med Image Anal 17: 365–374.

41. Handwerker DA, Ollinger JM, D’Esposito M (2004): Variation of BOLD hemodynamic responses across subjects and brain regions and their effects on statistical analyses. Neuroimage 21: 1639–1651.

42. David O, Guillemain I, Saillet S, Reyt S, Deransart C, Segebarth C, Depaulis A (2008): Identifying neural drivers with functional MRI: an electrophysiological validation. PLoS Biol 6: 2683–2697.

43. Xia M, Wang J, He Y (2013): BrainNet Viewer: a network visualization tool for human brain connectomics. PLoS One 8: e68910.

44. Pedrotti M, Lei S, Dzaack J, Rötting M (2011): A data-driven algorithm for offline pupil signal preprocessing and eyeblink detection in low-speed eye-tracking protocols. Behav Res Methods 43: 372–383.

45. Inhoff AW, Radach R (1998): Definition and computation of oculomotor measures in the study of cognitive processes. Eye Guidance in Reading and Scene Perception. Elsevier, pp 29–53.

46. Radach R, Heller D, Inhoff A (1999): Occurrence and function of very short fixation durations in reading. Current Oculomotor Research. Boston, MA: Springer US, pp 321–331.

47. Savitzky A, Golay MJE (1964): Smoothing and differentiation of data by simplified least squares procedures. Anal Chem 36: 1627–1639.

48. Mack DJ, Belfanti S, Schwarz U (2017): The effect of sampling rate and lowpass filters on saccades - A modeling approach. Behav Res Methods 49: 2146–2162.

49. Salvucci DD, Goldberg JH (2000): Identifying fixations and saccades in eye-tracking protocols. Proceedings of the Symposium on Eye Tracking Research & Applications - ETRA ’00. presented at the the symposium, New York, New York, USA: ACM Press. https://doi.org/10.1145/355017.355028

50. Toh WL, Castle DJ, Rossell SL (2015): Facial affect recognition in body dysmorphic disorder versus obsessive-compulsive disorder: An eye-tracking study. J Anxiety Disord 35: 49–59.

51. Toh WL, Castle DJ, Rossell SL (2017): How individuals with body dysmorphic disorder (BDD) process their own face: a quantitative and qualitative investigation based on an eye-tracking paradigm. Cogn Neuropsychiatry 22: 213–232.

52. Kollei I, Horndasch S, Erim Y, Martin A (2017): Visual selective attention in body dysmorphic disorder, bulimia nervosa and healthy controls. J Psychosom Res 92: 26–33.

53. Unema PJA, Pannasch S, Joos M, Velichkovsky BM (2005): Time course of information processing during scene perception: The relationship between saccade amplitude and fixation duration. Vis cogn 12: 473–494.

54. Burr DC, Morrone MC, Ross J (1994): Selective suppression of the magnocellular visual pathway during saccadic eye movements. Nature 371: 511–513.

55. Schall JD, Morel A, King DJ, Bullier J (1995): Topography of visual cortex connections with frontal eye field in macaque: convergence and segregation of processing streams. J Neurosci 15: 4464– 4487.

56. Antes JR (1974): The time course of picture viewing. J Exp Psychol 103: 62–70.

57. Over EAB, Hooge ITC, Vlaskamp BNS, Erkelens CJ (2007): Coarse-to-fine eye movement strategy in visual search. Vision Res 47: 2272–2280.

58. Velichkovsky BM (2002): Heterarchy of cognition: the depths and the highs of a framework for memory research. Memory 10: 405–419.

59. Wilhelm S, Phillips KA, Didie E, Buhlmann U, Greenberg JL, Fama JM, et al. (2014): Modular cognitive-behavioral therapy for body dysmorphic disorder: a randomized controlled trial. Behav Ther 45: 314–327.

60. Beilharz F, Castle DJ, Phillipou A, Rossell SL (2018): Visual training program for body dysmorphic disorder: protocol for a novel intervention pilot and feasibility trial. Pilot Feasibility Stud 4: 189.

61. Navon D (1977): Forest before trees: The precedence of global features in visual perception. Cogn Psychol 9: 353–383.

62. Beilharz FL, Atkins KJ, Duncum AJF, Mundy ME (2016): Altering visual perception abnormalities: A marker for body image concern. PLoS One 11: e0151933.

63. Taqui AM, Shaikh M, Gowani SA, Shahid F, Khan A, Tayyeb SM, et al. (2008): Body Dysmorphic Disorder: gender differences and prevalence in a Pakistani medical student population. BMC Psychiatry 8: 20.

64. Koran LM, Abujaoude E, Large MD, Serpe RT (2008): The prevalence of body dysmorphic disorder in the United States adult population. CNS Spectr 13: 316–322.

65. Bohon C, Hembacher E, Moller H, Moody TD, Feusner JD (2012): Nonlinear relationships between anxiety and visual processing of own and others’ faces in body dysmorphic disorder. Psychiatry Res 204: 132–139.

