## Supplementary Material for "Effects of visual attention modulation on dynamic effective connectivity and visual fixation during own-face viewing in body dysmorphic disorder"

**Title**

**Supplementary Methods**

1. Exclusion criteria
2. Clinical assessments
3. Image acquisition
4. Anatomical data preprocessing
5. Functional data preprocessing
6. Brain activation analysis
7. Brain connectivity analysis

**Supplementary Table**

**Table S1**

**Supplementary Figures**

**Figure S1**

**Figure S2**

**Figure S3**

**Figure S4**

**Supplementary References**

1. **Exclusion criteria**

The healthy controls were excluded if they had any current DSM-5 disorder or lifetime bipolar disorder or psychosis. Other exclusion criteria that applied to all participants included suicidality, self-injurious behavior, lifetime neurological disorder, current pregnancy, any medical illness that could affect cerebral metabolism, or current treatment with cognitive-behavioral therapy.

2. **Clinical assessments**

***Mini International Neuropsychiatric Interview (MINI):*** The MINI 7.0.0 is a short, structured diagnostic interview developed by psychiatrists and clinicians for DSM-5 and ICD-10 psychiatric disorders. With an administration time of ~15 minutes, the MINI is the structured psychiatric interview of choice for psychiatric evaluation and outcome tracking in clinical psychopharmacology trials and epidemiological studies. The MINI is the most widely used psychiatric structured diagnostic interview instrument in the world, employed by mental health professionals and health organizations in more than 100 countries.

***BDD Diagnostic Module (BDD-DM):*** The BDD-DM is a brief semi-structured diagnostic interview for BDD.

***Yale-Brown Obsessive-Compulsive Scale Modified for BDD (BDD-YBOCS):*** The BDD-YBOCS is a 12-item semi-structured, clinician-rated measure of current BDD severity used in many BDD studies. The first 5 items assess obsessional preoccupations about perceived appearance defects (time preoccupied, interference in functioning and distress due to perceived appearance defects, resistance against preoccupations, and control over preoccupations). Items 6-10 assess BDD-related repetitive behaviors and are similar to items 1-5 (time spent performing the behaviors, interference in functioning due to the behaviors, distress experienced if the behaviors are prevented, and resistance of and control over the behaviors). Item 11 assesses insight into appearance beliefs, and item 12 assesses avoidance because of BDD symptoms. Scores for each item range from 0 (no symptoms) to 4 (extreme symptoms); the total score ranges from 0 to 48, with higher scores reflecting more severe symptoms.

***Montgomery-Åsberg Depression Rating Scale (MADRS):*** The MADRS is a 10-item diagnostic questionnaire which psychiatrists used to measure the severity of depressive episodes. The questionnaire includes questions on the following symptoms: 1) apparent sadness, 2) reported sadness, 3) inner tension, 4) reduced sleep, 5) reduced appetite, 6) concentration difficulties, 7) lassitude, 8) inability to feel, 9) pessimistic thoughts, and 10) suicidal thoughts. Higher score indicates more severe depression, and each item yields a score of 0 to 6. The overall score ranges from 0 to 60.

***Brown Assessment of Beliefs Scale (BABS):*** The BABS is a 7-item semi-structured rater-administered scale that assesses insight/delusionality both dimensionally and categorically. BABS items assess the person’s conviction that their belief is accurate, perception of others’ views of the belief, explanation for any difference between the person’s and others’ views of the belief, whether the person could be convinced that the belief is wrong, attempts to disprove the belief, insight, and ideas/delusions of reference related to the belief. The first 6 items are summed to create a total score that ranges from 0 to 24; higher scores indicate poorer insight. Item 7 is not included in the total score, because referential thinking is characteristic of some but not all disorders.

***Hamilton Anxiety Scale (HAMA):*** The HAMA is a 14-item psychological questionnaire used by clinicians to rate the severity of a patient’s anxiety. Each of the 14 items is defined by a series of symptoms, and measures both psychic anxiety (mental agitation and psychological distress) and somatic anxiety (physical complaints related to anxiety). Each item is rated on a scale of 0 (not present) to 4 (severe), with a total score range of 0-56.

3. **Image acquisition**

A 3T Siemens Prisma scanner was used to obtain the MR images. A high-resolution T1-weighted structural image was acquired using ultrafast gradient echo sequence for anatomical reference (TR/TE: 2300/2.27 ms; flip angle: 8°; 256 x 256 matrix; voxel size: 1 x 0.977 x 0.977 mm3; 192 slices). Functional images were acquired using a T2*-weighted echo planar imaging (EPI) gradient-echo pulse sequence (TR/TE: 2500/25 ms; flip angle: 80°; 64 x 64 matrix; voxel size: 3 mm3, with a 0.75 mm gap; 34 slices; 124 volumes per run).

4. **Anatomical data preprocessing**

The T1-weighted (T1w) image was corrected for intensity non-uniformity (INU) (1) with ANTs 2.2.0 (2), and used as T1w-reference. The T1w-reference was skull-stripped, and brain tissue segmentation of cerebrospinal fluid (CSF), white-matter (WM) and gray-matter (GM) was performed on the brain-extracted T1w using FSL 5.0.9 (3). Brain surfaces were reconstructed using FreeSurfer 6.0.1 (4). Volume-based spatial normalization to standard space was performed through nonlinear registration with ANTs 2.2.0, using brain-extracted versions of both T1w reference and the T1w template (MNI152NLin2009cAsym) (5).

5. **Functional data preprocessing**

For each of the 3 fMRI runs per subject, the following preprocessing was performed. A deformation field to correct for susceptibility distortions was estimated based on fMRIPrep’s fieldmap-less approach. The deformation field is that resulting from co-registering the BOLD reference to the same-subject T1w-reference with its intensity inverted (6). Based on the estimated susceptibility distortion, an unwarped BOLD reference was calculated, and was co-registered to the T1w reference using FreeSurfer 6.0.1 which implements boundary-based registration (7). Co-registration was configured with nine degrees of freedom to account for distortions remaining in the BOLD reference. Head-motion parameters with respect to the BOLD reference were estimated before any spatiotemporal filtering using FSL 5.0.9 (8). The BOLD timeseries with slice-timing correction were resampled onto their native space by applying a single, composite transform to correct for head-motion and susceptibility distortions. The BOLD timeseries were resampled into standard MNI space, generating the spatially-normalized, preprocessed BOLD runs. Automatic removal of motion artifacts using independent component analysis (ICA-AROMA) (9) was performed on the preprocessed BOLD timeseries on MNI space after removal of non-steady state volumes and spatial smoothing with an isotropic, Gaussian kernel of 6mm FWHM. Corresponding “non-aggressively” denoised runs were produced after such smoothing. Several confounding timeseries were calculated based on the preprocessed BOLD: framewise displacement (FD), DVARS and three region-wise global signals. FD and DVARS were calculated for each run (10). The three global signals were extracted within the CSF, the WM, and the whole-brain masks. A set of physiological regressors were also extracted to allow for component-based noise correction (CompCor) (11). Principal components were estimated after high-pass filtering the preprocessed BOLD timeseries for the two CompCor variants: temporal (tCompCor) and anatomical (aCompCor). tCompCor components were calculated from the top 5% variable voxels within a mask covering the subcortical regions. aCompCor components were calculated within the intersection of the aforementioned mask and the union of CSF and WM masks calculated in T1w space. Components were also calculated separately within the WM and CSF masks. For each CompCor decomposition, the k components with the largest singular values were retained, such that the retained components’ timeseries were sufficient to explain 50% of variance across the nuisance mask. The remaining components were dropped from consideration. Frames that exceeded a threshold of 0.5 mm FD or 1.5 standardized DVARS were annotated as motion outliers.

6. **Brain activation analysis**

***First-level processing***

The ICA-AROMA preprocessed fMRI data in the MNI space were used for brain activation analysis. Brain extraction was done using *fslmaths* command with utilizing the brain mask in MNI space for the corresponding functional scan. Motion outliers were calculated using *fsl_motion_outliers* command on the brain-extracted functional scan to generate a motion outlier matrix. Moreover, a matrix of timepoints that fMRIPrep classified as non-steady state were extracted from the fMRIPrep confound file. These two matrices were concatenated in Matlab to generate the final outlier matrix which was then used as a confound EV matrix in FSL FEAT (12,13). FSL FEAT version 5.0.9 was run with the following settings: 10% for brain/background threshold, 0.66% for noise level, 0.34 for temporal smoothness, 5.3 for Z threshold, and 90s for high-pass filter cutoff. For Stats, FILM Pre-whitening was turned on, confound EVs and temporal derivatives were added, and temporal filtering was applied. For Post-Stats, cluster thresholding was used with Z threshold of 2.3, and p threshold of 0.05. Two contrasts were used: Faces > Baseline, and Baseline > Faces.

***Second-level processing***

After first-level processing was complete and outputs were visually inspected for errors, a second level of analysis was run to combine the three functional scans for each participant and create contrasts that were input into the third level of analysis. The fixed effects model had 3 explanatory variables: 1^st^ NatV, 2^nd^ NatV, and ModV. Four contrasts were used as inputs to higher level analysis: 1^st^ NatV > 2^nd^ NatV, 1^st^ NatV < 2^nd^ NatV, 1^st^ NatV > ModV, and 1^st^ NatV < ModV.

***Third-level processing***

After second-level processing was complete, a third level of analysis was conducted to investigate the group effects. The main hypotheses tested whether there were any within-group differences in BDD or CON for any of the lower-level contrasts between 1^st^ NatV and ModV, and between 1^st^ NatV and 2^nd^ NatV, and whether there were between-group differences in the observed effects. We expected that compared to NatV, the ModV would result in higher DVS activity and lower VVS activity when viewing faces vs. baseline. The difference between the conditions was expected to be larger in BDD than in controls. These hypotheses were tested by conducting the third level of analysis using the first-level contrast (faces > baseline) and the second-level contrasts (1^st^ NatV > 2^nd^ NatV, 1^st^ NatV < 2^nd^ NatV, 1^st^ NatV > ModV, and 1^st^ NatV < ModV). A 2-factor with 2-level ANOVA model was created for group and order. There were 4 EVs: BDD_NNM_, BDD_NMN_, CON_NNM_, and CON_NMN_. Preliminary F-tests using Flame1 were conducted for 3 contrasts: main group effect, main order effect, and group x order interaction. The final hypothesis testing model used the same EVs with 4 contrasts: BDD mean, CON mean, BDD > CON, and BDD < CON.

***Creation of DVS and VVS masks***

The DVS mask was created starting with the Harvard-Oxford atlas 10% thresholded visual stream mask which includes both dorsal and ventral visual streams. This mask subtracted the Harvard-Oxford atlas 10% thresholded 3-region VVS mask to get the DVS mask. The DVS mask was further polished by overlapping with a brain template to get rid of any non-brain areas.

The VVS mask was created by combining the Harvard-Oxford atlas 10% thresholded 3-region VVS mask and the Harvard-Oxford atlas 10% thresholded lateral occipital cortex. This VVS mask then subtracted the areas overlapped with the DVS mask.

7. **Brain connectivity analysis**

A similar analysis strategy has been adopted in our previous study for exploring dynamic connectivity for fear processing in BDD and Anorexia Nervosa (14). We used dynamic effective connectivity (DEC) in this study because of its ability to estimate causal connectivity across time with a precision of each time point, which helped us capture connectivity only within task blocks of interest. DEC was estimated in a Kalman filter based dynamic multivariate vector autoregressive model (dMVAR) framework that is based on the concept of Granger causality (GC). Simulations (15,16) as well as experimental results from electrophysiology and optogenetics (17–20) have demonstrated the ability of GC in estimating fMRI effective connectivity, when latent neural time series data are used after HRF deconvolution (as done in this study).

FMRI is an indirect measure of neural activity corrupted by blood hemodynamics that can alter the relative phase between fMRI time series without having any neural delays between the corresponding regions (21,22). Since this phenomenon can confound fMRI connectivity estimates (23), deconvolution was performed. Additionally, deconvolution has also been found to improve GC estimation (18,19,24). A blind-deconvolution technique developed by Wu et al. (25) was used, which is a data-driven technique based on the point process model (26). It identifies neural events in fMRI data as point processes based on relative intensities and estimates the best fit double-gamma HRF in a least squares sense. Latent neuronal time series is then estimated using Wiener deconvolution. Several studies have employed this technique, e.g., (27–29). The deconvolution was performed using rsHRF toolbox implemented in SPM12 (<https://github.com/compneuro-da/rsHRF>).

DEC was computed at each time point using Kalman-filter based time-varying GC (30). The basic concept is that if past values of a timeseries can forecast the future values of another timeseries, a causal influence from the former to the latter is inferred. A GC value of 0 represents no causal relationship from source to target timeseries, a value of 1 represents strong positive causality (i.e. increase in BOLD response of the source enhances BOLD response of the target, and vice versa), and a value of -1 represents strong negative causality (i.e. increase in BOLD response of the source suppresses BOLD response of the target, and vice versa) (31). The deconvolved timeseries were fitted into a dMVAR model for estimating DEC between ROIs, as in prior studies (32,33). Further details and underlying mathematics are elaborated in (31,34). The dMVAR model coefficients vary as a function of time, whose lengths were identical to the number of timepoints in the timeseries. That is, a DEC value was obtained for every timepoint for every connection. Using this, we obtained the desired block-specific connectivity values and aggregated them to represent the corresponding connection (35). The timepoints associated with those trials of viewing unaltered faces were extracted for subsequent statistical analysis.

***Repetition time issue in dynamic connectivity analysis***

A limitation in this study is the poor repetition time (TR) of 2.5s, which is barely sufficient to capture effective connectivity in the brain (36). With this modest TR, the data is not sensitive enough to capture causal interactions happening at faster timescales. In other words, a shorter TR would have enabled us to capture more DEC patterns than with a longer TR (or a longer TR could only capture a subset of DEC patterns compared with the patterns captured with a shorter TR). It is possible that there are within-group and/or between-group differences not detected with the existing TR of 2.5s, leading to false negative errors (where the test results incorrectly fail to indicate the presence of a condition when it is present). But importantly, the positive results of this study are still reliable as it does not imply that the positive results captured at a longer TR are less valid compared to what would have been captured at a shorter TR. These aspects must be kept in mind while interpreting negative findings of this study. Future studies must prioritize faster acquisitions in their study designs and tradeoffs so that maximum benefit is drawn from dynamic time series analysis techniques (such as those used in this study).

**Table S1** Sample characteristics

| **Group** | **Order** | **Sex (Male/Female)** | **Age (Years)** | **Symptoms Severity** | | | |
| --- | --- | --- | --- | --- | --- | --- | --- |
|  |  |  |  | **HAMA** | **MADRS** | **BDD-YBOCS** | **BABS** |
| BDD (n=37) | NNM (n=18) | 2/16 | 26.1±8.2 | 10.7±6.2 | 14.2±9.8 | 27.1±5.0 | 15.4±4.4 |
|  | NMN (n=19) | 4/15 | 23.6±5.0 | 9.3±8.1 | 10.2±8.2 | 26.5±3.5 | 15.0±4.7 |
| CON (n=30) | NNM (n=16) | 6/10 | 23.3±6.4 | 3.1±2.3 | 1.2±1.4 | NA | NA |
|  | NMN (n=14) | 2/12 | 23.1±7.5 | 1.9±2.2 | 1.0±1.3 | NA | NA |
| Chi-Square Tests | χ^2^ (order=NNM) | 3.278 |  |  |  |  |  |
|  | P-value (order=NNM) | 0.070 |  |  |  |  |  |
|  | χ^2^ (order=NMN) | 0.248 |  |  |  |  |  |
|  | P-value (order=NMN) | 0.618 |  |  |  |  |  |
| Tests of Between-Subjects Effects | F  (group*order) |  | 0.469 | 0.010 | 1.269 | NA | NA |
|  | P-value (group*order) |  | 0.496 | 0.922 | 0.264 | NA | NA |
|  | F  (order) |  | 0.626 | 0.850 | 1.534 | 0.142 | 0.109 |
|  | P-value (order) |  | 0.432 | 0.360 | 0.220 | 0.708 | 0.744 |
|  | F  (group) |  | 1.037 | 29.454 | 43.996 | NA | NA |
|  | P-value (group) |  | 0.313 | <0.001 | <0.001 | NA | NA |

BDD = body dysmorphic disorder; CON = control; HAMA = Hamilton Anxiety Scale; MADRS = Montgomery-Åsberg Depression Rating Scale; BDD-YBOCS = Yale-Brown Obsessive-Compulsive Scale Modified for BDD; BABS = Brown Assessment of Beliefs Scale; χ^2^ = chi-square test; F = F-test from two-way ANOVA.

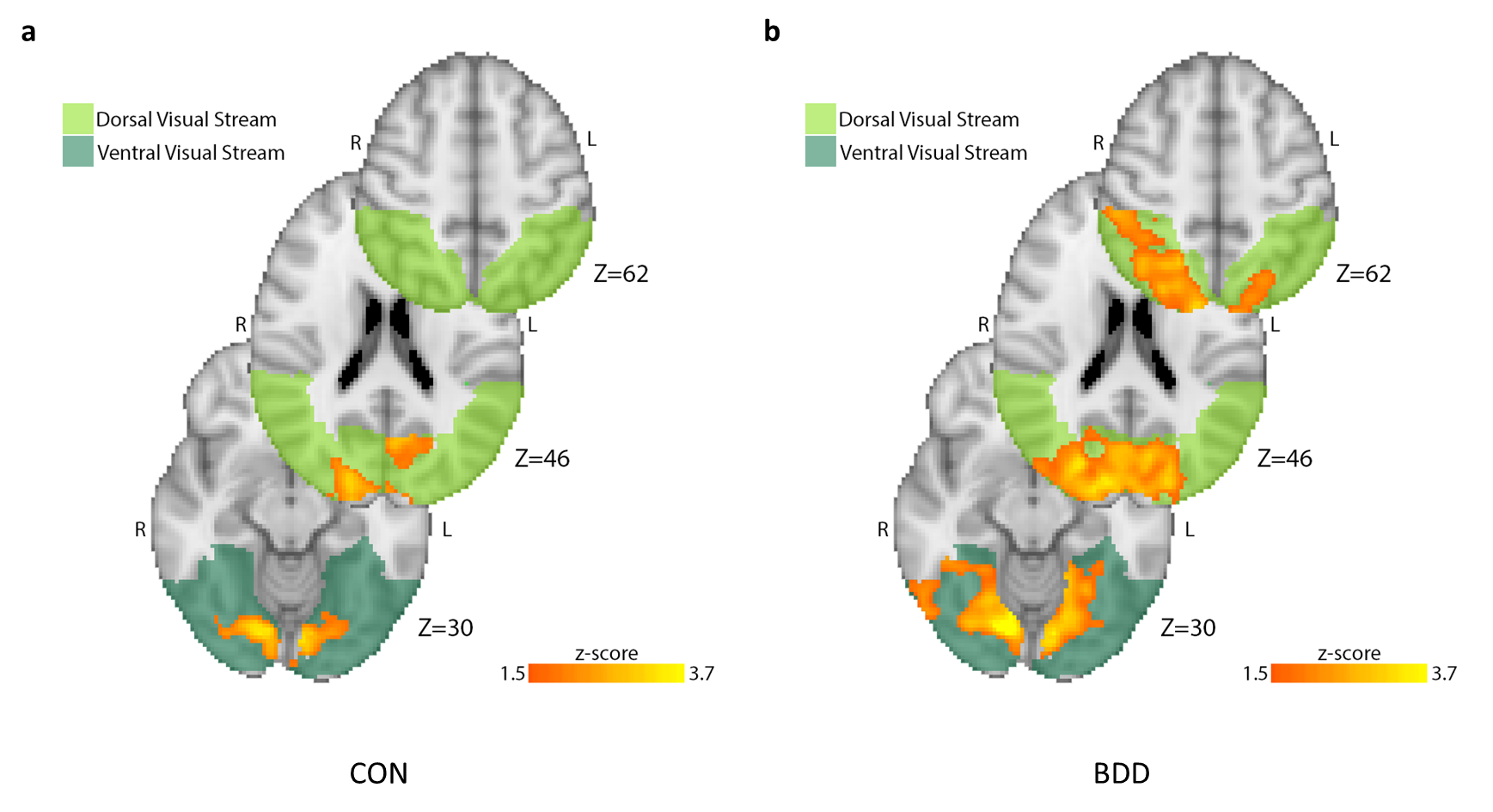

**Figure S1** Brain activation maps obtained from higher-level analysis for (a) CON and (b) BDD groups during face viewing. The activation in the DVS and VVS in response to unaltered face stimuli relative to baseline was greater during 1^st^ NatV compared to ModV for both CON and BDD. The significant cluster was larger in BDD compared to CON. Activated clusters were significant at p<0.05 corrected for multiple comparisons. The z coordinate of each image is indicated in the axial views of the activation maps.

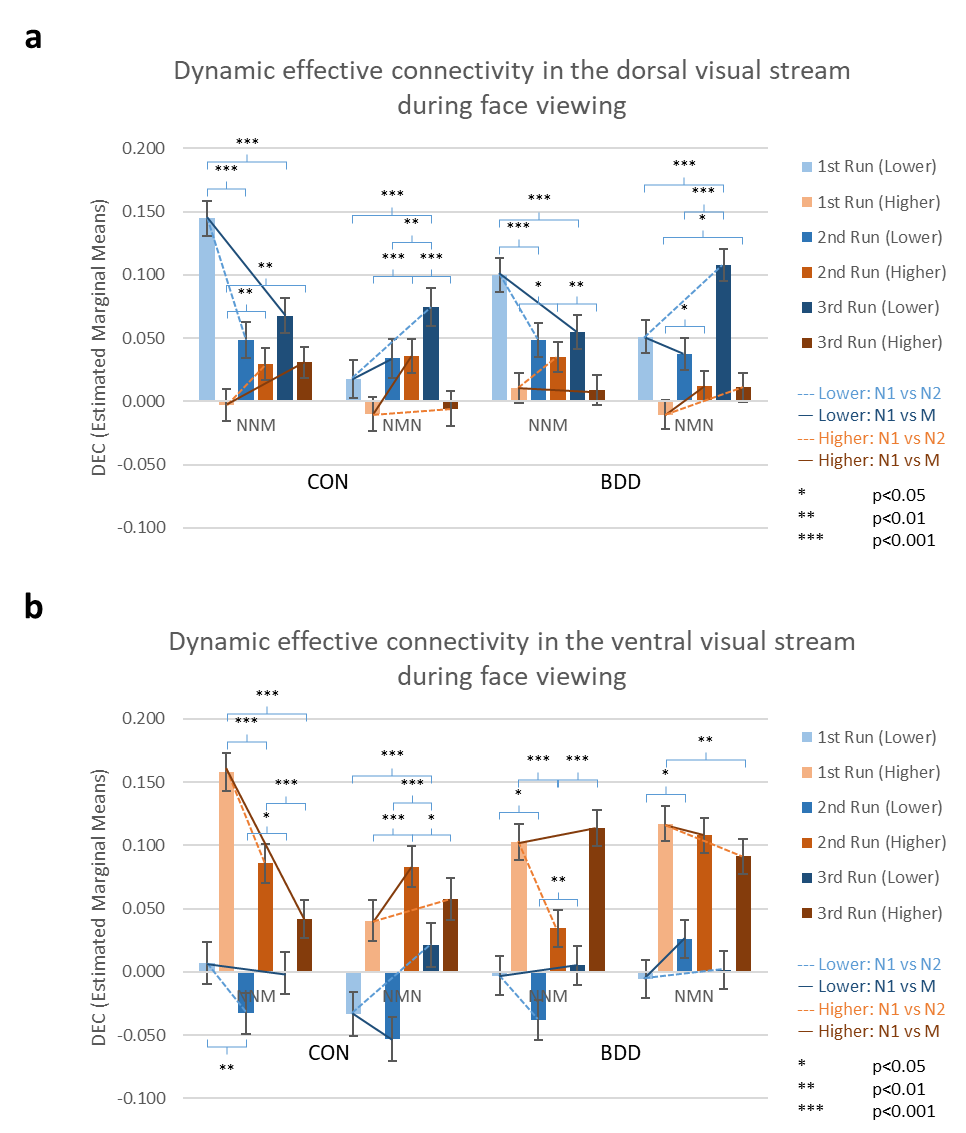

**Figure S2** Estimated marginal means of dynamic effective connectivity for the (a) dorsal visual stream and (b) ventral visual stream across groups, orders and runs. The CON and BDD groups with NNM and NMN orders represent the two counterbalanced orders for each group, with the corresponding means for the 1^st^, 2^nd^ and 3^rd^ runs indicated in the bar plots. For example, the healthy controls randomized to CON with NNM received natural viewing (N), natural viewing (N), and then modulated viewing (M) as the 1^st^, 2^nd^ and 3^rd^ runs.

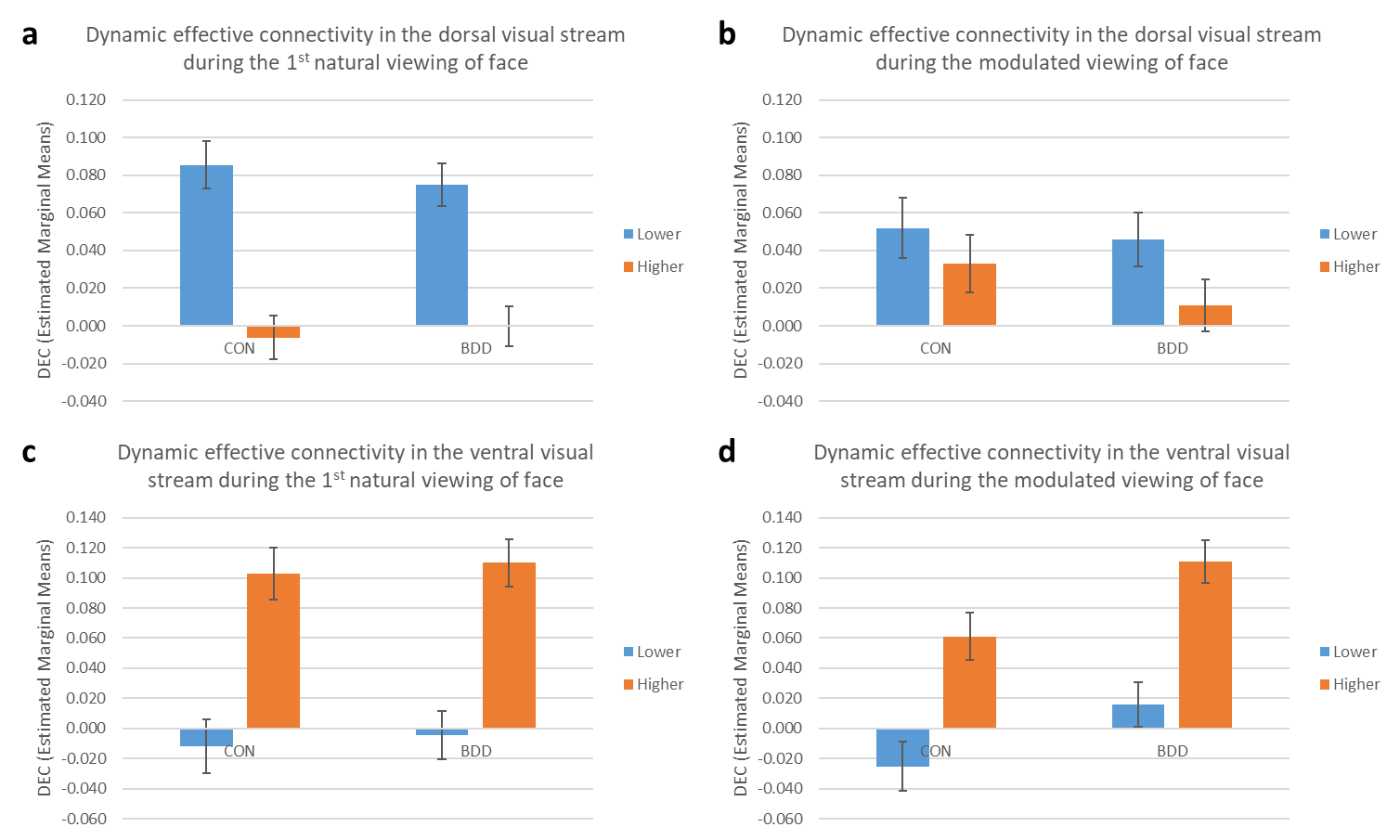

**Figure S3** Estimated marginal means of dynamic effective connectivity for the (a, b) dorsal visual stream and (c, d) ventral visual stream across CON and BDD groups during the first natural viewing and modulated viewing of face, with collapsing the ‘order’ and ‘run’ effects. In the DVS during both the first natural viewing and modulated viewing, there was significant effect for ‘level’ (p<0.001). In the VVS during the first natural viewing, there was significant effect for ‘level’ (p<0.001). In the VVS during the modulated viewing, there were significant effects for ‘group’ (p=0.033) and ‘level’ (p<0.001).

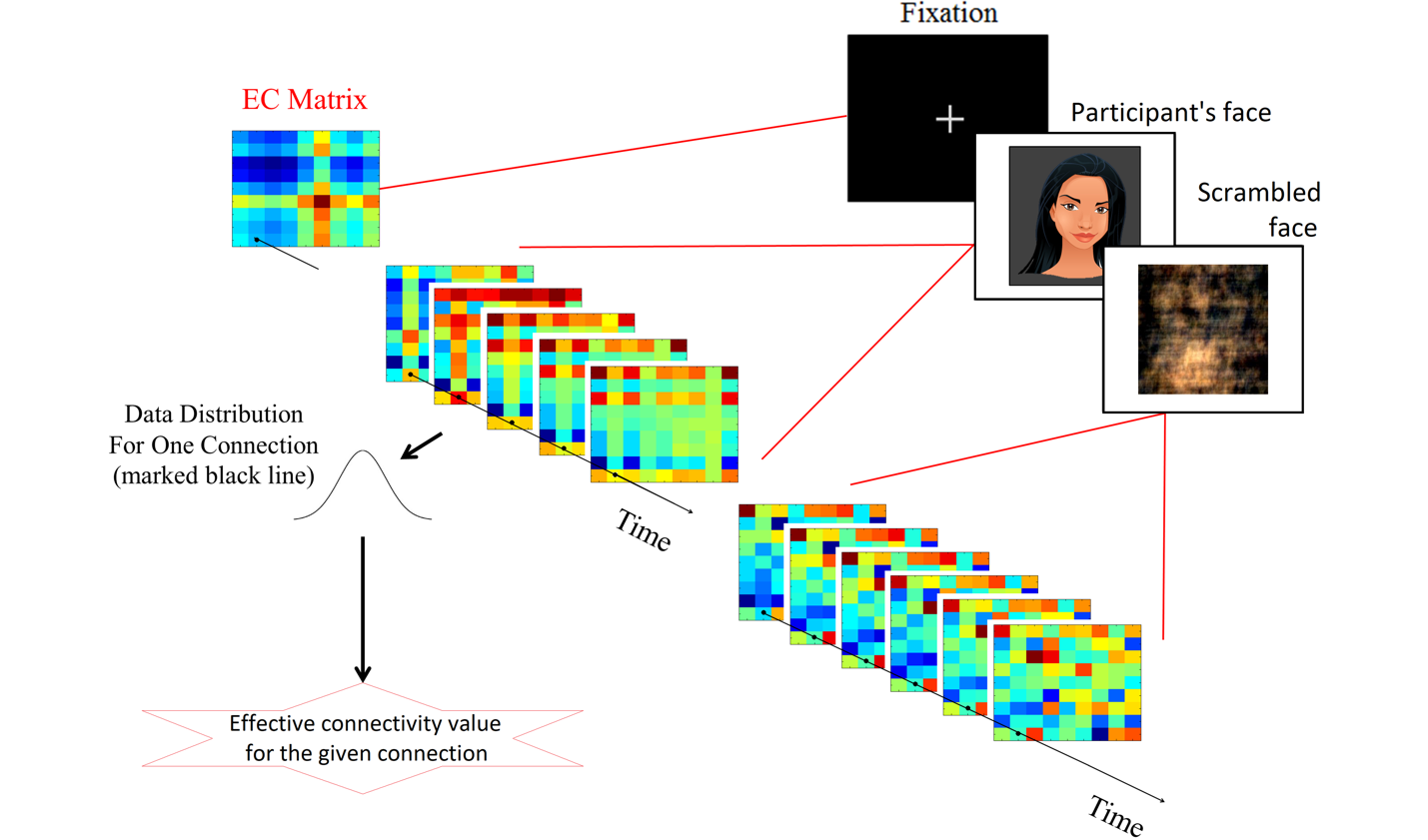

**Figure S4** A simplistic demonstration of dynamic effective connectivity analysis to estimate the directional connectivity value from our task fMRI data. Effective connectivity (EC) matrices, estimated for each time point, were pooled across task blocks of viewing one's own face to derive our final EC estimate for each connection.

**References**

1. Tustison NJ, Avants BB, Cook PA, Zheng Y, Egan A, Yushkevich PA, Gee JC (2010): N4ITK: Improved N3 Bias Correction. *IEEE Transactions on Medical Imaging*, vol. 29. pp 1310–1320.

2. Avants B, Epstein C, Grossman M, Gee J (2008): Symmetric diffeomorphic image registration with cross-correlation: Evaluating automated labeling of elderly and neurodegenerative brain. *Medical Image Analysis*, vol. 12. pp 26–41.

3. Zhang Y, Brady M, Smith S (2001): Segmentation of brain MR images through a hidden Markov random field model and the expectation-maximization algorithm. *IEEE Transactions on Medical Imaging*, vol. 20. pp 45–57.

4. Dale AM, Fischl B, Sereno MI (1999): Cortical surface-based analysis. I. Segmentation and surface reconstruction. *Neuroimage* 9: 179–194.

5. Fonov VS, Evans AC, McKinstry RC, Almli CR, Collins DL (2009): Unbiased nonlinear average age-appropriate brain templates from birth to adulthood. *NeuroImage*, vol. 47. p S102.

6. Wang S, Peterson DJ, Gatenby JC, Li W, Grabowski TJ, Madhyastha TM (2017): Evaluation of Field Map and Nonlinear Registration Methods for Correction of Susceptibility Artifacts in Diffusion MRI. *Frontiers in Neuroinformatics*, vol. 11. https://doi.org/10.3389/fninf.2017.00017

7. Greve DN, Fischl B (2009): Accurate and robust brain image alignment using boundary-based registration. *NeuroImage*, vol. 48. pp 63–72.

8. Jenkinson M, Bannister P, Brady M, Smith S (2002): Improved Optimization for the Robust and Accurate Linear Registration and Motion Correction of Brain Images. *NeuroImage*, vol. 17. pp 825–841.

9. Pruim RHR, Mennes M, van Rooij D, Llera A, Buitelaar JK, Beckmann CF (2015): ICA-AROMA: A robust ICA-based strategy for removing motion artifacts from fMRI data. *NeuroImage*, vol. 112. pp 267–277.

10. Power JD, Mitra A, Laumann TO, Snyder AZ, Schlaggar BL, Petersen SE (2014): Methods to detect, characterize, and remove motion artifact in resting state fMRI. *NeuroImage*, vol. 84. pp 320–341.

11. Behzadi Y, Restom K, Liau J, Liu TT (2007): A component based noise correction method (CompCor) for BOLD and perfusion based fMRI. *NeuroImage*, vol. 37. pp 90–101.

12. Woolrich MW, Ripley BD, Brady M, Smith SM (2001): Temporal autocorrelation in univariate linear modeling of FMRI data. *Neuroimage* 14: 1370–1386.

13. Woolrich MW, Behrens TEJ, Beckmann CF, Jenkinson M, Smith SM (2004): Multilevel linear modelling for FMRI group analysis using Bayesian inference. *Neuroimage* 21: 1732–1747.

14. Rangaprakash D, Bohon C, Lawrence KE, Moody T, Morfini F, Khalsa SS, *et al.* (2018): Aberrant dynamic connectivity for fear processing in anorexia nervosa and body dysmorphic disorder. *Front Psychiatry* 9: 273.

15. Wen X, Rangarajan G, Ding M (2013): Is Granger causality a viable technique for analyzing fMRI data? *PLoS One* 8: e67428.

16. Ryali S, Supekar K, Chen T, Menon V (2011): Multivariate dynamical systems models for estimating causal interactions in fMRI. *Neuroimage* 54: 807–823.

17. Katwal SB, Gore JC, Gatenby JC, Rogers BP (2013): Measuring relative timings of brain activities using fMRI. *Neuroimage* 66: 436–448.

18. David O, Guillemain I, Saillet S, Reyt S, Deransart C, Segebarth C, Depaulis A (2008): Identifying neural drivers with functional MRI: an electrophysiological validation. *PLoS Biol* 6: 2683–2697.

19. Ryali S, Shih Y-YI, Chen T, Kochalka J, Albaugh D, Fang Z, *et al.* (2016): Combining optogenetic stimulation and fMRI to validate a multivariate dynamical systems model for estimating causal brain interactions. *Neuroimage* 132: 398–405.

20. Wang Y, Katwal S, Rogers B, Gore J, Deshpande G (2017): Experimental validation of dynamic Granger causality for inferring stimulus-evoked sub-100 ms timing differences from fMRI. *IEEE Trans Neural Syst Rehabil Eng* 25: 539–546.

21. Aguirre GK, Zarahn E, D’esposito M (1998): The variability of human, BOLD hemodynamic responses. *Neuroimage* 8: 360–369.

22. Handwerker DA, Ollinger JM, D’Esposito M (2004): Variation of BOLD hemodynamic responses across subjects and brain regions and their effects on statistical analyses. *Neuroimage* 21: 1639–1651.

23. Rangaprakash D, Wu G-R, Marinazzo D, Hu X, Deshpande G (2018): Hemodynamic response function (HRF) variability confounds resting-state fMRI functional connectivity. *Magn Reson Med* 80: 1697–1713.

24. Ryali S, Chen T, Supekar K, Menon V (2012): Estimation of functional connectivity in fMRI data using stability selection-based sparse partial correlation with elastic net penalty. *Neuroimage* 59: 3852–3861.

25. Wu G-R, Liao W, Stramaglia S, Ding J-R, Chen H, Marinazzo D (2013): A blind deconvolution approach to recover effective connectivity brain networks from resting state fMRI data. *Med Image Anal* 17: 365–374.

26. Saad ZS, Gotts SJ, Murphy K, Chen G, Jo HJ, Martin A, Cox RW (2012): Trouble at rest: how correlation patterns and group differences become distorted after global signal regression. *Brain Connect* 2: 25–32.

27. Boly M, Sasai S, Gosseries O, Oizumi M, Casali A, Massimini M, Tononi G (2015): Stimulus set meaningfulness and neurophysiological differentiation: a functional magnetic resonance imaging study. *PLoS One* 10: e0125337.

28. Amico E, Gomez F, Di Perri C, Vanhaudenhuyse A, Lesenfants D, Boveroux P, *et al.* (2014): Posterior cingulate cortex-related co-activation patterns: a resting state FMRI study in propofol-induced loss of consciousness. *PLoS One* 9: e100012.

29. Lamichhane B, Adhikari BM, Brosnan SF, Dhamala M (2014): The neural basis of perceived unfairness in economic exchanges. *Brain Connect* 4: 619–630.

30. Büchel C, Friston KJ (1998): Dynamic changes in effective connectivity characterized by variable parameter regression and Kalman filtering. *Hum Brain Mapp* 6: 403–408.

31. Rangaprakash D, Dretsch MN, Venkataraman A, Katz JS, Denney TS Jr, Deshpande G (2018): Identifying disease foci from static and dynamic effective connectivity networks: Illustration in soldiers with trauma. *Hum Brain Mapp* 39: 264–287.

32. Hutcheson NL, Sreenivasan KR, Deshpande G, Reid MA, Hadley J, White DM, *et al.* (2015): Effective connectivity during episodic memory retrieval in schizophrenia participants before and after antipsychotic medication. *Hum Brain Mapp* 36: 1442–1457.

33. Feng C, Deshpande G, Liu C, Gu R, Luo Y-J, Krueger F (2016): Diffusion of responsibility attenuates altruistic punishment: A functional magnetic resonance imaging effective connectivity study. *Hum Brain Mapp* 37: 663–677.

34. Rangaprakash D, Dretsch MN, Katz JS, Denney TS Jr, Deshpande G (2019): Dynamics of segregation and integration in directional brain networks: Illustration in soldiers with PTSD and neurotrauma. *Front Neurosci* 13: 803.

35. Sathian K, Deshpande G, Stilla R (2013): Neural changes with tactile learning reflect decision-level reweighting of perceptual readout. *J Neurosci* 33: 5387–5398.

36. Abler B, Roebroeck A, Goebel R, Höse A, Schönfeldt-Lecuona C, Hole G, Walter H (2006): Investigating directed influences between activated brain areas in a motor-response task using fMRI. *Magn Reson Imaging* 24: 181–185.
